# SARS-CoV-2 diagnostic testing rates determine the sensitivity of genomic surveillance programs

**DOI:** 10.1101/2022.05.20.22275319

**Authors:** Alvin X. Han, Amy Toporowski, Jilian A. Sacks, Mark D. Perkins, Sylvie Briand, Maria van Kerkhove, Emma Hannay, Sergio Carmona, Bill Rodriguez, Edyth Parker, Brooke E. Nichols, Colin A. Russell

## Abstract

The first step in SARS-CoV-2 genomic surveillance is testing to identify infected people. However, global testing rates are falling as we emerge from the acute health emergency and remain low in many low- and middle-income countries (LMICs) (mean = 27 tests/100,000 people/day). We simulated COVID-19 epidemics in a prototypical LMIC to investigate how testing rates, sampling strategies, and sequencing proportions jointly impact surveillance outcomes and showed that low testing rates and spatiotemporal biases delay time-to-detection of new variants by weeks-to-months and can lead to unreliable estimates of variant prevalence even when the proportion of samples sequenced is increased. Accordingly, investments in wider access to diagnostics to support testing rates of ∼100 tests/100,000 people/day could enable more timely detection of new variants and reliable estimates of variant prevalence. The performance of global SARS-CoV-2 genomic surveillance programs is fundamentally limited by access to diagnostic testing.

## Introduction

Since the start of the COVID-19 pandemic in 2019, unprecedented expansion of genomic surveillance efforts has led to the generation of more than 10 million SARS-CoV-2 sequences deposited in the publicly accessible GISAID database (https://www.gisaid.org/) as of May 2022. These efforts have been integral to understanding the COVID-19 pandemic^1^, including the identification of the Alpha variant in the United Kingdom during the fall 2020^2^, the Delta variant in India in late 2020^3^, and the Omicron variant in Southern Africa in November 2021^4^. Despite the value of these efforts for monitoring the evolution of SARS-CoV-2, the intensity of genomic surveillance is highly heterogenous across countries. High-income countries (HICs) on average produced 16 times more SARS-CoV-2 sequences per reported case than low- and middle-income countries (LMICs) as a result of longstanding socioeconomic inequalities and consequent underfunding of laboratory and surveillance infrastructures^5^. To strengthen global pandemic preparedness, initiatives such as the Access to COVID-19 Tools Accelerator Global Risk Monitoring Framework, the Pan American Health Organization COIVD-19 Genomic Surveillance Regional Network, the Africa Pathogen Genomics Initiative, as well as the Global Influenza Surveillance and Response System, among others, have supported LMICs in developing pathogen genomic surveillance programs.

As resources are finite, it is critical that sequencing sample sizes, and the diagnostic testing needed to obtain samples for sequencing, are carefully set for genomic surveillance programs to detect and monitor variants as efficiently as possible. Current recommended sample sizes are based on sampling theory^5–8^ and assume that the volume of diagnostic testing is large enough such that the diversity of sampled viruses is representative of the diversity of viruses circulating in the population. However, LMICs test at a mean rate of 27 tests per 100,000 persons per day (tests/100k/day) as opposed to >800 tests/100k/day across HICs based on observational data collected between January 2020 and March 2022^9^, with even higher testing rates in some HICs (Fig. 1). Low testing rates lead to spotty information and smaller virus specimen pools available for sequencing, resulting in sampling biases. These factors can render efforts to monitor the emergence of new variants or prevalence of existing variants highly unreliable.

**Fig. 1.**
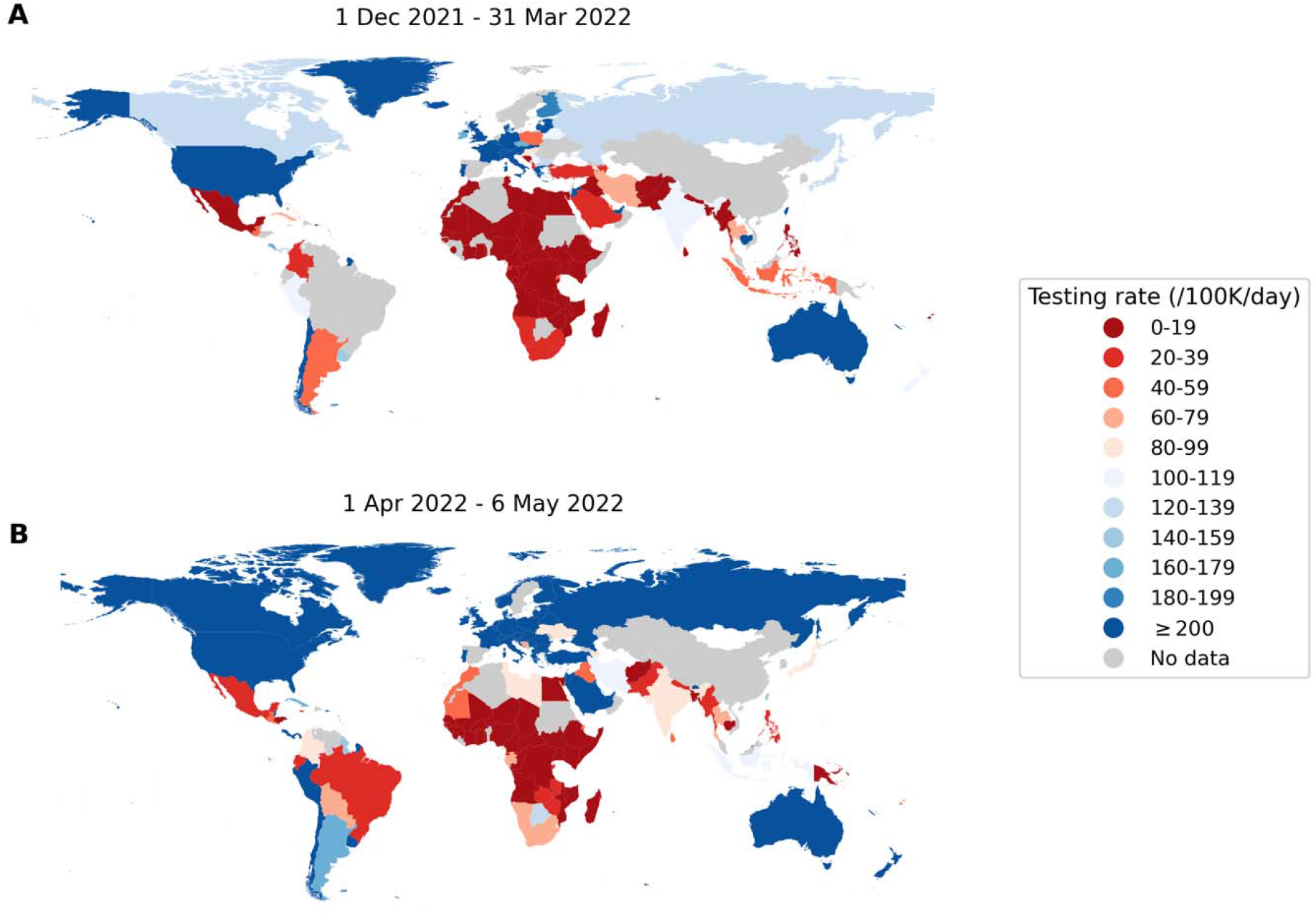
Global disparities in SARS-CoV-2 testing rates. Each country is colored by the average total number of SARS-CoV-2 tests performed per 100,000 persons per day (/100K/day) (**A**) between 1 December 2021 and 31 March 2022 when the Omicron variant-of-concern spread around the world; (**B**) between 1 April 2022 and 6 May 2022 when most countries were past peak Omicron wave of infections^9^.

Here, we studied how different testing rates can impact genomic surveillance outcomes. Specifically, we developed and used the *Propelling Action for Testing And Treating* (PATAT) model, an individual-based modelling framework, to simulate concurrently-circulating wild-type SARS-CoV-2 (pre-Alpha viruses)/Alpha-like epidemics as well as Delta-/Omicron (BA.1)-like epidemics in Zambia as a representative LMIC archetype where recent demographic census data required by the model was available (Online Methods). We assumed that Alpha and Omicron (BA.1) were more transmissible than the respective extant virus to achieve growth rates of ∼0.15/day and ∼0.35/day respectively^2,10^ and simulated SARS-CoV-2 infection waves in a population of 1,000,000 individuals over a 90-day period that begins with an initial 1% prevalence of the extant SARS-CoV-2 variant and the mutant variant being introduced at 0.01%. We assumed that clinic-based professional-use Antigen Rapid Diagnostic Tests (Ag-RDTs) form the basis of testing given persistent reports that polymerase chain reaction (PCR) tests are poorly accessible for detection of symptomatic cases outside of tertiary medical facilities in many LMICs^11^.

We then simulated different genomic surveillance sampling strategies to elucidate how testing, sequencing volumes and the degree of sampling bias arising from sources of specimens jointly impact the timeliness of variant detection and the accuracy of variant monitoring (Online Methods). These strategies include: (i) sending all samples from community clinics and tertiary hospitals to a centralized facility for possible sequencing (i.e. population-wide strategy); (ii) sampling and sequencing a portion of positive specimens collected at one tertiary sentinel facility for the population of 1,000,000 simulated people (including mild individuals seeking symptomatic testing and severe patients who sought tertiary care at the facility); or sampling and sequencing a portion of positive specimens collected at (iii) 10%, (iv) 25%, (v) 50%, and (vi) 100% of all tertiary sentinel facilities.

## Results

### Performance of current guidance

We first assessed various suggested sample sizes of positive specimens to sequence to detect SARS-CoV-2 variants at low prevalence for simulated wild-type/Alpha and Delta/Omicron epidemics in Zambia with a mean testing rate of 27 tests/100k/day (based on the observed mean rate of testing in LMICs) (Fig. 2). We used recommended sample sizes from three prominent guidances: (1) The World Health Organization and European Centre for Disease Prevention and Control computed sample size using the binomial method^7,8^; (2) By subsampling genomic surveillance data generated in Denmark in 2020-2021 when the country was testing at >2,000 tests/100k/day on average, Brito et al. suggested that sequencing 0.5% of all detected cases with a turnaround time of 21 days would result in a 20% of variant detection before reaching 100 cases^5^; (3) Wohl et al. formulated a novel framework computing sequencing sample size by modeling the biological and logistical processes that impact sampled variant proportions^6^. Critically, all three methods did not consider how low testing rates and spatial nonuniformity in sampling coverage impact sampled variant proportions, and in turn, speed of variant detection. The assumptions, mathematical background, and lack of accounting for spatiotemporal bias in sample size estimation of each guidance are detailed in Table 1 and Supplementary Notes.

**Fig. 2.**
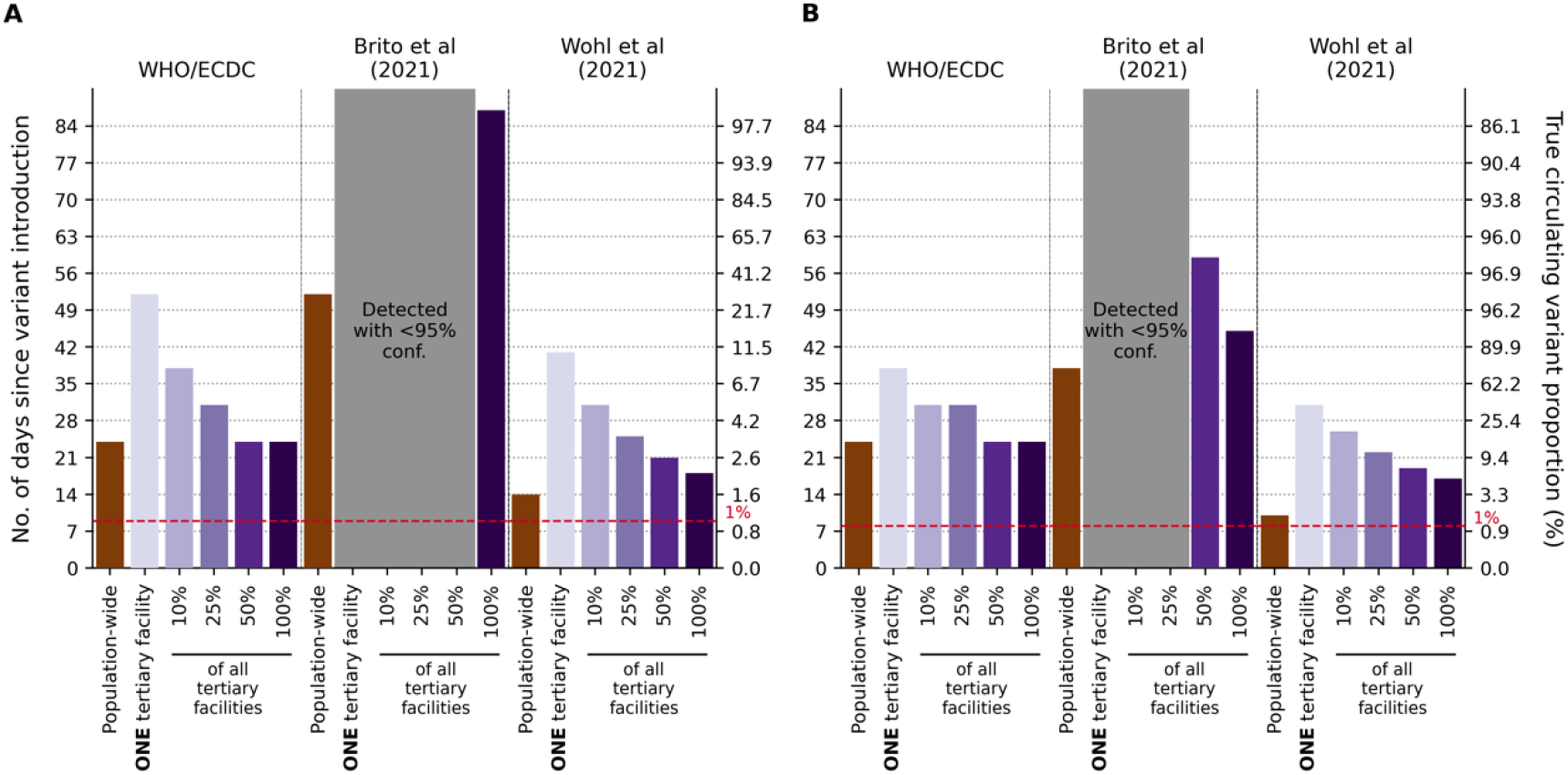
Performance of current guidance on number of positive specimens to sequence for variant detection with testing rate at 27 tests per 100,000 persons per day. First day of detection since variant introduction at 95% confidence and the corresponding circulating variant proportion using guidance from the World Health Organization (WHO)/European Centre for Disease Prevention and Control (ECDC) ^7,8^, Brito et al. ^5^, and Wohl et al. ^6^ (Table 1) under different genomic surveillance strategies with varying sampling coverage (i.e. all collected specimens from all healthcare facilities are sent to one facility to be sampled for sequencing (*population-wide* strategy); only *one*, 10%, 25%, 50%, or 100% of tertiary sentinel facilities would sample the specimens they collected for sequencing). Turnaround time (i.e. time from specimen collection to acquisition of sequencing data) was assumed to be negligible. 1,000 random independent simulations were performed for each guidance/surveillance strategy. We simulated epidemics for (**A**) Wild-type SARS-CoV-2/Alpha. (**B**) Delta/Omicron. Grey regions denote that we could not reliably detect the variant virus with 95% confidence using the guidance in question under the assumed genomic surveillance strategy.

**Table 1.** Current guidance by various stakeholder and academic groups on the number of specimens to sequence for detection of novel variants at low prevalence.

|  | Recommendation on number/proportion of positive cases to sequence |  | Critical considerations |
| --- | --- | --- | --- |
| World Health Organization (WHO)/European Centre for Disease Prevention and Control (ECDC)<sup>7,8</sup> | *No. of positive cases* | *No. of sequences to detect at 1% with 95% confidence* | • Agnostic to variant properties<br>• Assumes specimen pool to be sampled for sequencing is representative of circulating diversity but acknowledges that unless testing coverage is evenly distributed this will be a biased sample<br>• Notes that in countries with limited sequencing capacity monitoring relative prevalence of variants should be prioritized |
|  | <1000 cases | 141 |  |
|  | 1001 – 2,500 | 196 |  |
|  | 2,500 – 5,000 | 243 |  |
|  | 5,001 – 10,000 | 270 |  |
|  | >10,000 | 285 |  |
| Brito et al., 2021<sup>5</sup> | At least 0.5% of all cases with a turnaround time of 21 days to detect novel lineage before it reaches 100 cases at 20% probability |  | • Based on sequencing data from Denmark which is testing at an average of >2,000 tests per 100,000 persons per day<sup>9</sup> |
| Wohl et al., 2022<sup>6</sup> | 1-29 sequences per day to detect an Alpha-like variant based on 0.03% initial introduction for a population of 10,000 (assuming growth rate of 0.1/day) at 1% with 95% confidence.<br>We used the spreadsheet (https://github.com/HopkinsIDD/VOCsamplesize) provided and input appropriate parameters to obtain the recommendation relevant to the simulated epidemics. |  | • Assumes that the *observed* variant proportion in the positive specimens collected is representative of the *circulating* variant proportions among the infected population. This requires a large number of specimens that are randomly collected for assumption to hold true at low circulating variant proportions<br>• A correction factor is included to correct for biases in the *observed* variant proportion but only pertaining to those arising from the relative differences in diagnostic sensitivity, sample qualities and conditional asymptomatic and symptomatic testing probabilities between the two circulating variants. |

As such, even when assuming negligible turnaround time (i.e. time from specimen collection to acquisition of sequencing data), the recommended approaches were insufficient to detect the variant on their respective target detection day when testing rates were low, due to poor representativeness, regardless of the genomic surveillance sampling strategy. The first strategy of sampling specimens collected from the whole population that were sent to one sequencing facility (i.e. population-wide strategy) led to the best performance (closest to target detection day) for all recommendations, as it involves random uniform sampling of all available samples, a fundamental assumption made by all current guidance. However, if the specimen pools available for sequencing are restricted to those collected from a subset of sentinel tertiary facilities only, the non-uniformity in sampling coverage results in spatiotemporal bias within the sequenced samples, and leads to delayed detection of variants-of-concern (VOCs), which gets progressively worse as the proportion of tertiary facilities performing sequencing decreases to one facility.

### Variant detection

To elucidate how SARS-CoV-2 testing rates and the proportion of positive specimens sequenced impact the speed of variant detection, we simulated wild-type SARS-CoV-2/Alpha and Delta/Omicron epidemics at different Ag-RDT availability ranging from 27 tests/100k/day to 1,000 tests/100k/day (Fig. 3). We assumed that specimens to be sequenced are sampled on their collection day, and varied the proportion of positive specimens to sample for sequencing each day between 1% and 100%. We analyzed the impact of testing rates and sequencing proportions on the expected day when the first specimen sampled for sequencing containing the variant was collected as a measure of variant detection speed. In Fig. 3, we did not consider the time between sample collection and sequencing nor the turnaround time to obtaining sequencing results as they would only delay the actual day of variant detection by the assumed turnaround time.

**Fig. 3.**
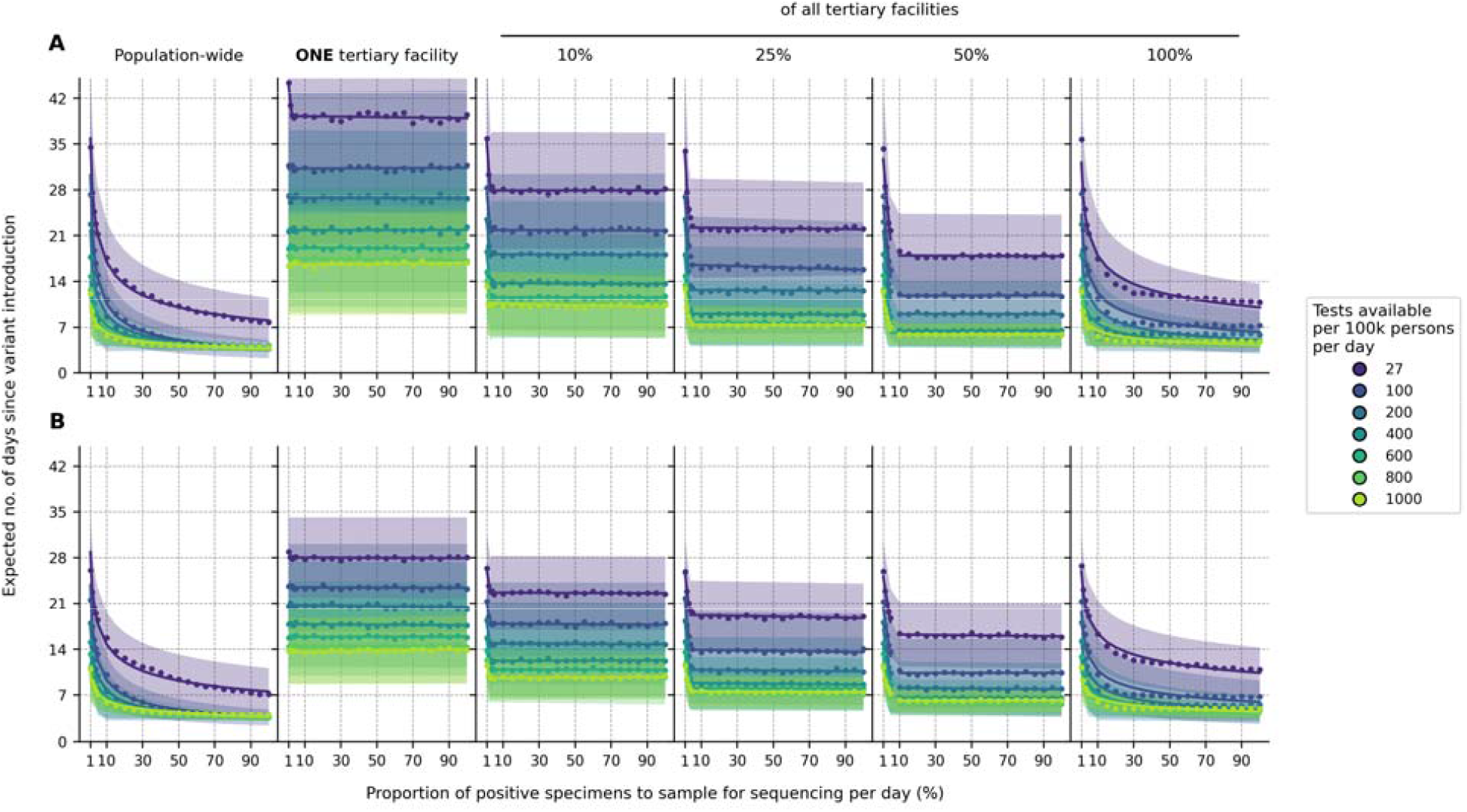
Impact of SARS-CoV-2 testing rates and proportion of positive specimens to sequence on variant detection. For each mean daily test availability (line and shading color), the expected day when the first variant specimen to be sequenced is sampled since its introduction is plotted against the proportion of positive specimens to be sampled for sequencing daily. Different genomic surveillance strategies with varying sampling coverage (i.e. all specimens collected from all healthcare facilities sent to one facility to be sampled for sequencing (population-wide strategy); only one, 10%, 25%, 50%, or 100% of tertiary sentinel facilities would sample the specimens they collected for sequencing) were simulated. (**A**) Wild-type SARS-CoV-2/Alpha. (**B**) Delta/Omicron. The expected day when the first variant specimen is sampled was computed from 1,000 random independent simulations for each surveillance strategy. The shaded region depicts the standard deviation across simulations.

For all testing rates, the relationship between the expected day when the first sample containing the variant was collected and the proportion of positive specimens sequenced per day can be described by a convex operating curve, reflecting rapidly diminishing returns in the speed of variant detection as more specimens are sampled for sequencing. Across all genomic surveillance sampling strategies, relatively larger marginal improvements to the speed of variant detection are generally made when the sequencing proportion is increased up to ∼10% of all samples collected. Further sequencing only minimally shortens the expected time to variant detection, as the operating curve asymptotically approaches the earliest possible day of detection. Importantly, increasing SARS-CoV-2 testing allows smaller sequencing proportions to attain similar detection day targets, and higher testing rates lower the earliest possible detection day. For both the Alpha and Omicron variants, increasing testing rates from 27 tests/100k/day to 100 tests/100k/day brings forward the expected day of sampling the first variant sequence by at least one week (Fig. 3).

For the same level of testing and sequencing proportion, the population-wide strategy led to the earliest initial detection of a variant sequence. If sequencing were restricted to samples collected at a subset of tertiary sentinel facilities only, increasing the number of facilities sending samples for sequencing reduced the spatiotemporal bias in the specimen pool, thereby shaping the operating curves closer to the ones observed for the population-wide strategy. Interestingly, results similar to the population-wide strategy could be attained if all tertiary facilities acted as sentinel sites and sent the samples they collected for sequencing to increase the representativeness of sampling.

### Observed variant proportion

Test availability and sampling coverage also affect the accuracy of the observed variant proportion (Figs. 4 and Extended Data S1). At a testing rate of 27 tests/100k/day, the observed variant proportion maximally differs from the true circulating proportion by >30% for both the Alpha and Omicron variants and for more than 15% of the time, the proportional difference between the observed and true variation was greater than 20%. Both the maximum absolute difference and percentage of timepoints where the difference is >20% can be lowered to <20% and <5% respectively if testing rate is increased to 100 or more tests/100k/day.

**Fig. 4.**
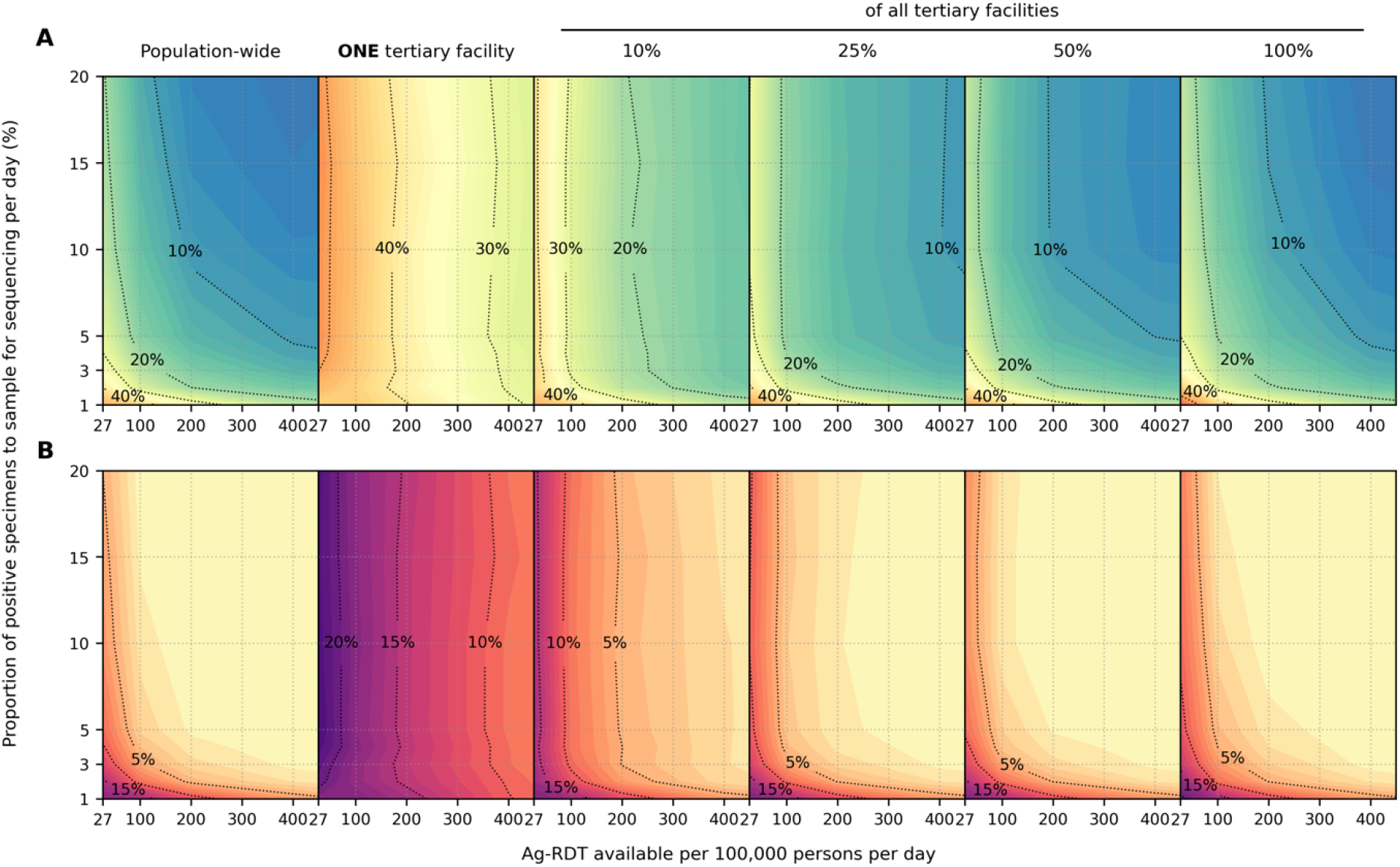
Impact of SARS-CoV-2 testing rates on the capacity to monitor changes in variant prevalence based on diagnostic test availability and proportion of test-positive samples sequenced. Different genomic surveillance strategies (i.e. all specimens collected from all healthcare facilities sent to one facility to be sampled for sequencing (*population-wide* strategy); only *one*, 10%, 25%, 50%, or 100% of tertiary sentinel facilities would sample the specimens they collected for sequencing) were simulated. (**A**) Maximum absolute difference between observed and circulating variant proportions. (**B**) Proportion of timepoints when sequencing was performed that the absolute difference between observed and circulating variant proportions is greater than 20%. All results were computed from 1,000 random independent simulations for each surveillance strategy.

Critically, when the representativeness of the specimen pool is spatiotemporally biased by sequencing samples collected at tertiary sentinel facilities only, increasing the proportion of specimens to be sequenced only marginally lowers the maximum absolute difference or lessens the number of times where observed variant proportion deviates less than 20% from true circulating proportions (Fig. 4, near vertical isoclines at low daily rates of testing). Increasing testing rates at sentinel surveillance sites provides more accurate detection in changes to circulating prevalence than sequencing more samples in the context of low testing rates.

### Sensitivity analyses

We repeated our analyses using virus properties (i.e., incubation period, maximum viral load, protection against infection by the mutant virus after extant virus infection) of the Omicron variant but varied different relative transmissibility to the Delta variant (1.0 to 4.0) as well as the initial proportion of individuals who had been infected by the Delta variant (10% and 40%). The variant growth rates simulated for these hypothetical Delta/Omicron epidemics ranged from 0.17/day to 0.42/day.

Under these varied conditions, the expected day when the specimen of the first variant sequence is collected still follows a convex-shaped operating curve against the daily proportion of positive specimens to sequence. For all curves, the larger marginal improvements in shortening variant detection are still in sequencing proportions of up to ∼10% (Extended Data Fig. S2). In terms of the accuracy of observed variant to true circulating proportions, the maximum absolute difference and percentage of timepoints where difference is >20% are both substantially lowered if testing rate is increased to at least 100 tests/100k/day (Extended Data Fig. S3-4).

We also varied the prevalence of extant Delta infections when the Omicron variant was introduced (Extended Data Fig. S5). We found that lower test availability causes a delay in sampling the first variant specimen if the variant is introduced when pre-existing extant variant circulation is high. At 27 tests/100k/day, regardless of specimen proportions sequenced, detection could be delayed by ∼1 week if Omicron was introduced when Delta was circulating at 10% prevalence as opposed to 1%. This is because a greater share of tests would be used to diagnose the more prevalent extant virus infections which in turn decreases the likelihood of detecting the newly introduced variant at low proportions.

## Discussion

Our findings show that the emphasis on the proportion of samples referred for genomic surveillance is misplaced if testing capacity is insufficient and sample sources are highly spatiotemporally biased. As such, at the current mean rate of testing in LMICs (27 tests/100k/day), current guidance^5–8^ on sequencing sample size estimation could likely lead to later-than-predicted detection of novel variants at best or, at worst, leave new variants undetected until they have infected a majority of a population.

Based on our work, we identified three major areas of improvement that could be prioritized to enhance the robustness of genomic surveillance programs (Fig. 5). First, the most substantial improvements are likely to come from increasing the mean testing rate in LMICs from 27 tests/100k/day (Fig. 5A) to at least 100 tests/100k/day (Fig. 5B). Even if one were to conduct sentinel surveillance only at one tertiary facility, this increase in testing rate for the catchment area of the facility would speed up variant detection by 1-2 weeks.

**Fig. 5.**
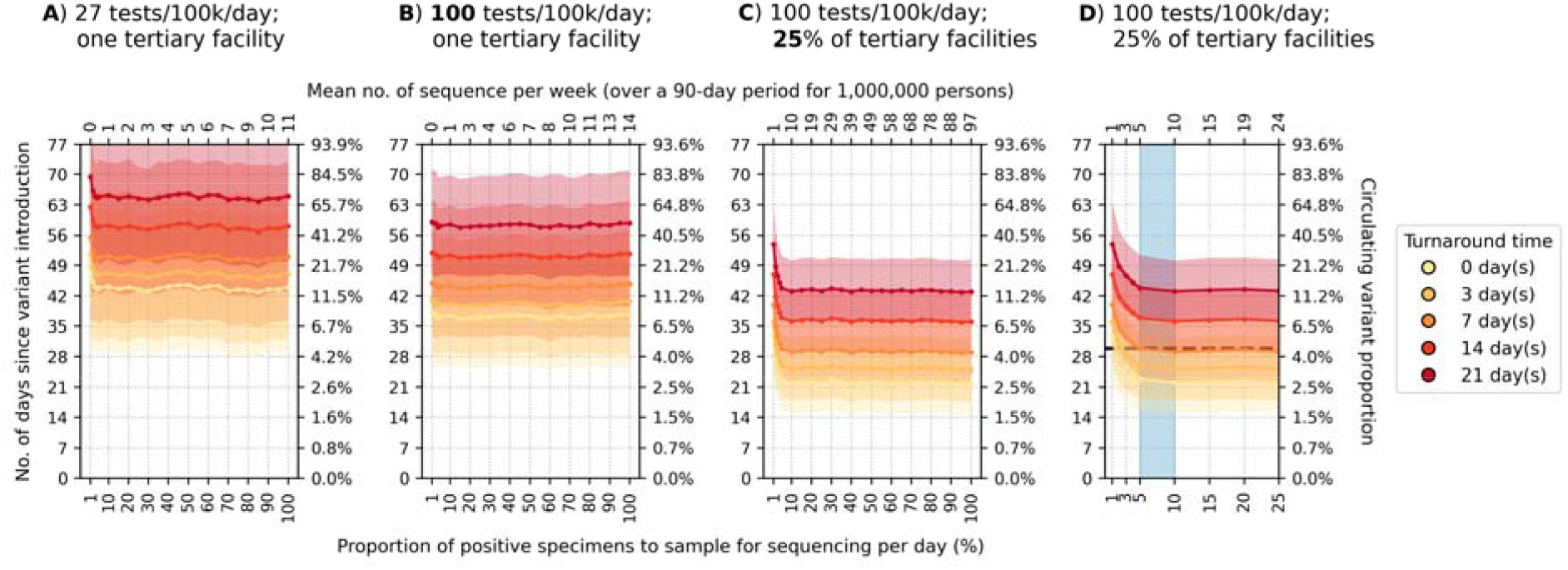
Recommended approach to enhance genomic surveillance robustness. In each plot, the operating curves of the expected day when the first Alpha variant sequence is generated are plotted for different proportion of specimens to sample for sequencing per day and turnaround times. We assumed that the Alpha variant was circulating at 1% initially with wild-type SARS-CoV-2 in the background. We also assumed that positive specimens sampled within each week for sequencing are consolidated into a batch before they are referred for sequencing. Turnaround time refers to the time between collection of each weekly consolidated batch of positive specimens to the acquisition of its corresponding sequencing data. The vertical axes denote the number of days passed since the introduction of the Alpha variant (left) and its corresponding circulating proportion (right). The horizontal axes denote the proportion of positive specimens to sample for sequencing per day (bottom) and the corresponding mean number of sequences to be generated per week per 1,000,000 people over a 90-day epidemic period. (**A**) Specimen pools for sequencing from *one* tertiary sentinel facility with testing rate at 27 tests per 100,000 persons per day (tests/100k/day). (**B**) Specimen pools for sequencing from *one* tertiary sentinel facility with testing rate at 100 tests/100k/day. (**C**) Specimen pools for sequencing from 25% of all tertiary sentinel facilities with testing rate at 100 tests/100k/day. (**D**) Zoomed-in plot of (C) to highlight sequencing proportions varying between 1-25%. Sequencing 5-10% of positive specimens (blue shaded region) would ensure that we would expectedly detect Alpha in 30 days if turnaround time is kept within one week. All results were computed from 1,000 random independent simulations for each surveillance strategy. The shaded region depicts the standard deviation across simulations.

Second, the representativeness of a specimen pool for sequencing can be further improved by expanding sampling coverage. In our model, variant detection was further sped up by 1-3 weeks by increasing the percentage of tertiary sentinel facilities sending the samples they had collected for sequencing to 25% of facilities (Fig. 5C). Additionally, in terms of prevalence monitoring, if 25% of tertiary facilities sequenced 5% of all positive specimens they had collected to detect and monitor an Alpha-like variant, the maximum absolute difference to true circulating proportion is expected to decrease from >50% (assuming a single sentinel facility) to no more than 20%.

Third, reducing turnaround time from samples referred to sequencing output results in a 1:1 decrease in time to new variant detection regardless of the proportion of sequenced samples, test availability or sampling coverage (Fig. 5). These gains require scale up in sample transport networks, access to sequencing machinery, trained personnel, and/or increases in numbers of sequenced samples to make the most efficient use of each sequencing run^11^. Furthermore, LMICs also often face high costs and extended delivery delays of laboratory reagents and consumables that were sometimes further exacerbated by recurring travel bans during the acute phase of the pandemic^5,12,13^.

After reducing spatiotemporal bias in the specimen pool through increased testing and sampling coverage, sequencing up to 5-10% of the positive specimens collected could return the greatest information gains while minimizing resource wastage. For an Alpha-like variant, 100 tests/100k/day with sampling from 25% of tertiary sentinel facilities for sequencing amounts to an estimated 5-10 sequences per week averaged over a 90-day period per 1,000,000 people. If turnaround time is kept within one week, the variant would likely be detected within one month at ∼4% circulating proportion (Fig. 5D). Similarly, at the same testing rate, sampling coverage and turnaround time (i.e. average 5-11 sequences per week per 1,000,000 people), an Omicron-like variant would be detected before the first month since its introduction but at ∼23% circulating proportion owing to its faster transmission (Extended Data Fig. S6).

Our findings serve to inform expectations of genomic surveillance initiative and should be interpreted according to the public health objectives of each program. If the objective is to serve as an early warning system for the *de novo* emergence of new variants before they are likely to have spread widely, then all factors above can be considered essential and could require substantially more than 100 tests/100k/day. Critically, determining that a new variant is a threat requires not only detection of the variant itself but also the capacity to reliably monitor changes in its prevalence and potential clinical impact on short timescales. The results presented here also inform the design of programs for the sensitive and reliable detection of changes in variant prevalence. Otherwise, if the objective is to detect for introduction of novel variants from overseas, some of the factors above may be relaxed depending on the public health objectives.For instance, if the aim is to attempt containment, all factors should still be considered to promptly detect and monitor the spread of the variant. However, if the aim to ensure sufficient time for control strategies to be enacted, less samples could be sequenced or turnaround time could be longer, for example, so long as the mitigation strategies remain useful when implemented.

Despite performing our simulations using demographic parameters from Zambia, the emergence and detection of each VOC to date represents interesting case studies for the work described here (Supplementary Notes). For example, at the time of first detection of the Omicron variant, in South Africa in November 2021, the daily SARS-CoV-2 testing rate was 51 tests/100,000 people/day^9^, which was among the highest testing rates in Africa. However, the Omicron variant was only detected 6-8 weeks after its likely emergence^4^. At that point, Omicron had already infected a substantial portion of the population in Gauteng, South Africa (i.e., the estimated circulating variant proportion was >80% by mid-November)^4^. Not only had the variant already spread across the rest of South Africa and to neighboring Botswana^4^, Omicron samples were also collected in multiple other countries, including Hong Kong^14^, Denmark^15^, and the Netherlands^16^ before the initial reports on the identification of the Omicron variant. This situation is consistent with our modelling findings, where novel variant detection is possible with <100 tests/100k/day but only after the new variant has spread widely across the population.

In another example, Germany randomly sequenced ∼60-70 sequences per week (i.e. <1% of cases sequenced per day) in December 2020^17^. During this time, testing rates in Germany averaged at ∼300 tests/100k/day^9^. Germany was able to detect the Alpha variant one week before WHO declared the lineage a variant-of-concern in mid-December 2020^17^. The Alpha variant likely emerged in the UK in mid-September 2020^18^ and rapidly proliferated across the country before it was reported in December 2020^19^. Our analyses showed that the expected time before the first Alpha variant specimen was sampled for sequencing since its introduction is >4 weeks (i.e. around November 2020) at Germany’s testing and sequencing rate. This falls in line with the likely period of Alpha’s introduction into Germany, similar to the period estimated for its European neighbors such as the Netherlands^20^.

While we find that routine representative sampling is vital for monitoring SARS-CoV-2 evolution, additional surveillance systems, including targeted surveillance of particular populations and settings (such as immunocompromised individuals or unusual events), and wastewater sampling, could enable increased variant detection sensitivity^21^. In particular, recent advances in wastewater sequencing and deconvolution methods to resolve multiple viral lineages in mixed wastewater samples enabled detection of emerging variants before they were captured by clinical genomic surveillance^22–24^. However, sequence quality is often poor in wastewater samples and in turn, these methods depend on a priori knowledge of the lineage-defining mutations of VOCs and variants-of-interest, which are currently still identified based on significant upsurges in clinically diagnosed cases. Furthermore, centralized wastewater management systems, which these methods rely on for accurate determination of relative lineage prevalence, are currently non-existent in many LMICs. Substantial investments, coordination and time are needed to enable local sanitation infrastructures suitable for wastewater surveillance^25^.Detection of genetic markers such as S-gene target failure in PCR assays may also provide faster notification of viral lineages with these specific mutations. However, whole genome sequencing is still needed for unambiguous genotyping of SARS-CoV-2 samples. Ultimately, clinical diagnostic testing and surveillance will remain the core mode of SARS-CoV-2 surveillance in most countries.

During the initial phase of the pandemic in 2020, due to limited testing and sequencing capacities, many LMICs were initially focused on genomic surveillance efforts at points of entry at country borders to deter introductions^26–28^. Over time, especially after the emergence of VOCs, SARS-CoV-2 genomic surveillance gradually expanded to include community surveillance as many LMICs enhanced their sequencing capacities^4,28–30^. This was done either by establishing regional sequencing networks to maximize available resources, investing in local sequencing capacities or partnering with global collaborators^30–32^. Sequencing turnaround time has also improved from an average of ∼170 days in 2020 to ∼30 days in 2021 across the African continent, albeit with substantial variation among countries^30^. While sequencing capabilities have expanded in LMICs, obtaining spatiotemporally representative samples remains a key challenge^30^. Our work shows that the sensitivity of genomic surveillance programs is highly dependent on diagnostic testing rate and that a mean testing rate of 100 tests/100k/day at sentinel sites that are geographically spread out across the community is a good basis for monitoring virus variants. While a reflexive PCR test after a positive Ag-RDT diagnosis is currently performed to obtain samples suitable for sequencing (and is possible in many tertiary facilities in LMICs), this presents additional cost and logistical barriers. Recent studies showed that SARS-CoV-2 sequencing can be performed using materials obtained from Ag-RDTs performed at point-of-care^33–35^. Importantly, whole genomes can be recovered up to eight days after testing, providing opportunities for sequencing to be performed on samples performed through self-testing as well^33^.

Expanding genomic sequencing capabilities, especially in LMICs, is a global priority^36^ and current investments in sequencing must continue^30,31^. Simultaneously, sustained investments in public health systems are required to expand access to, and availability of, diagnostic testing to underpin SARS-CoV-2 surveillance programs. Here, we primarily focused on LMICs but our findings on the impact of testing rates and representativeness on genomic surveillance programs are equally important for HICs as parts of their testing and surveillance infrastructures are dismantled following the acute health emergency of the COVID-19 pandemic. Ultimately, detecting the next SARS-CoV-2 variant or pathogen that causes the next pandemic requires fundamental clinical diagnostic capacity to monitor existing and emerging pathogens.

## Online Methods

### Simulating SARS-CoV-2 epidemics with the **P**ropelling **A**ction for **T**esting **A**nd **T**reating (PATAT) model

We used PATAT, a stochastic individual-based model to simulate SARS-CoV-2 epidemics in a community with demographic profiles, contact mixing patterns, and level of public health resources mirroring those typically observed in LMICs. Here, the model was based on Zambia. PATAT creates an age-structured population, linking individuals within contact networks of multi-generational households, schools, workplaces, and churches (i.e., regular mass gatherings) (Extended Data Table S1). The simulated number of healthcare facilities (i.e., community clinics and tertiary hospitals) where individuals with mild symptoms seek symptomatic testing and have their virus specimens collected was based on an empirical clinic-to-population ratio (i.e. one healthcare facility for every 7,000 individuals on average)^37,38^. Although PATAT does not explicitly simulate the spatial location of individuals, contact networks and healthcare facilities are ordered to approximate localized community structures (i.e. the closer the number order of a facility, the closer they are in the same neighborhood) that is most illustrative of urban centers. Households are proximally ordered and distributed around these facilities based on an empirical distance-structured distribution that correlates with probabilities of symptomatic individuals seeking testing at clinics (Extended Data Table S1).

We then simulated SARS-CoV-2 infection waves in a population of 1,000,000 individuals over a 90-day period that begins with an initial 1% prevalence of an extant SARS-CoV-2 variant and the introduction of a mutant variant at 0.01%. We assumed that clinic-based professional-use Antigen Rapid Diagnostic Tests (Ag-RDTs) are the predominant SARS-CoV-2 diagnostic used for SARS-CoV-2 testing. ^39^As Ag-RDT sensitivity depends on within-host viral loads^40^, PATAT generates viral load trajectories, measured in cycle threshold (Ct) values, for infected individuals by randomly sampling from known viral load distributions of different SARS-CoV-2 variants^41,42^. We performed simulations for two variant replacement scenarios – Alpha variant introduction while the wild-type virus was circulating (wild-type/Alpha) and Omicron (BA.1) variant introduction while Delta was circulating (Delta/Omicron), applying known distributions of their peak viral load, incubation, and virus clearance periods^41,43^ (Extended Data Table S1). Before simulating the two-variant epidemic, we first calibrated the transmission probability parameter for the extant variant such that it would spread in a completely susceptible population at R_0_ = 2.5-3.0. We then assumed Alpha and Omicron (BA.1) were more transmissible than the respective extant virus to achieve growth rates of ∼0.15/day and ∼0.35/day respectively^2,10^.

For both sets of simulations, we assumed that 10% of the population had infection-acquired immunity against the extant strain initially with some level of protection against infection by the mutant virus (wild-type SARS-CoV-2: 80% protection against Alpha^44^; Delta: 20% protection against Omicron^10^). We also investigated the scenario where 40% of the population had infection-acquired immunity as part of sensitivity analyses (see below). We did not investigate scenarios involving vaccine-acquired immunity due to low vaccine uptake in most LMICs^45^. PATAT uses the SEIRD (Susceptible-Exposed-Infected-Recovered/Death) epidemic model for disease progression and stratifies infected individuals based on their symptom presentation (asymptomatic, mild, or severe). After an assumed random delay post-symptom onset (mean = 1 day; s.d. = 0.5 day), symptomatic individuals who seek testing would do so at their nearest healthcare facility, where test-positive samples may be reflexively collected for sequencing. We assumed that symptomatic individuals sought testing based on a probability distribution of health services-seeking behaviour that inversely correlates with the distance between the individual’s household and the nearest healthcare facility (Extended Data Table S1)^46^.

We varied levels of Ag-RDT stocks per day (i.e., 27, 100, and 200-1,000 (in increments of 200) tests/100k/day), running 10 independent epidemic simulations for each testing rate. Given the start of a week on Monday, we assumed that a week’s worth of tests are delivered to healthcare facilities every Monday and unused Ag-RDTs in the previous week are carried forward into the next week. If test stocks for a particular week were exhausted before the end of the week, testing for the rest of that week ceased. Due to overlapping symptoms between COVID-19 and other respiratory diseases, a proportion of available Ag-RDTs would be used by individuals who are not infected with SARS-CoV-2. Based on test positivity rates reported by various countries in the second half of 2021^47^, we assumed 10% test positivity rate at the start and end of the simulated epidemic, and 20% test positivity at its peak, linearly interpolating the rates between these timepoints. We also assumed that false positive specimens could be sampled based on reported Ag-RDT specificity of 98.9%^40^.

We assumed that any specimens collected for genomic surveillance after positive detection through Ag-RDT would be reflexively confirmed with PCR. We also assumed that all symptomatic individuals who have severe symptoms require hospitalization, and are tested separately from mild symptomatic persons who sought testing. Given that likely only ∼10-20% of COVID-19 deaths in Zambia were tested for the disease in life^48,49^, we assumed that only 20% of individuals with severe disease would be tested by Ag-RDT or PCR upon presenting severe symptoms and have specimens collected for sequencing.

Full technical details of PATAT is described in the Supplementary Notes. The full model source code is available at https://github.com/AMC-LAEB/PATAT-sim.

### Genomic surveillance strategies

Twenty percent of healthcare facilities were assumed to be tertiary facilities based on empirical data collected from Zambia^37,38^. We assumed that tertiary facilities provide testing for mild symptomatic individuals as well as hospitalized patients with severe symptoms. Given that healthcare facilities were proximally ordered, we randomly selected tertiary facilities in each independent surveillance simulation (see below) but ensured that the selected facilities were not consecutively ordered. In sum, all tertiary facilities accounted for a median 18.4% (interquartile range = 17.7-19.1%) of total testing volume across all simulations. We assumed that a proportion of tertiary facilities serve as sentinel surveillance sites that reflexively collect SARS-CoV-2 positive samples for sequencing. We then simulated six strategies with varying degrees of sampling coverage where positive specimens collected from testing sites would be consolidated and sampled for sequencing: (i) all samples from community clinics and tertiary hospitals are sent to a centralized facility and further sampled for sequencing (i.e. *population-wide* strategy); (ii) only *one* tertiary sentinel facility for the population of 1,000,000 simulated people would sequence a portion of positive specimens it has collected, both from mild individuals seeking symptomatic testing and severe patients who sought tertiary care at the facility; or only (iii) 10%, (iv) 25%, (v) 50%, and (vi) 100% of all tertiary sentinel facilities would sample and sequence a proportion of the specimens they have collected.

For all strategies, we assumed that a proportion (1%-100%; in 2% increments between 1% and 5%, in 5% increments between 5% and 100%) of positive specimens are collected daily for sequencing. We also assumed that positive specimens sampled within each week for sequencing are consolidated into a batch before they are referred for sequencing. Turnaround time refers to the time between collection of each weekly consolidated batch of positive specimens to the acquisition of its corresponding sequencing data. Since the within-host viral loads of infected individuals were simulated, we assumed that only high-quality samples where Ct values < 30 could be sequenced and that sequencing success rate is 80% as assumed in other studies^6^.

For each strategy and sequencing proportion, we performed 100 independent surveillance simulations for each epidemic simulation with a given test stock availability, thus totaling to 1,000 random simulations for each set of variables (i.e., testing rate, sequencing proportion, and strategy).

## Data Availability

All data relevant to the study are included in the Article, the Supplementary Information and the GitHub repository (https://github.com/AMC-LAEB/PATAT-sim). The PATAT model source code is also available at https://github.com/AMC-LAEB/PATAT-sim.

## Acknowledgments

The authors are pleased to acknowledge that all computational work reported in this paper was performed on the Shared Computing Cluster which is administered by Boston University’s Research Computing Services (www.bu.edu/tech/support/research/). This work was supported by the Rockefeller Foundation, and the Governments of Germany, Canada, UK, Australia, Norway, Saudi Arabia, Kuwait, Netherlands and Portugal. A.X.H. and C.A.R. were supported by ERC NaviFlu (No. 818353). C.A.R. was also supported by NIH R01 (5R01AI132362-04) and an NWO Vici Award (09150182010027).

## Author contributions

Conceptualization: A.X.H., B.E.N., C.A.R., J.A.S., M.D.P., S.B., M.V.K., A.T., E.H., S.C., B.R.; Methodology: A.X.H., B.E.N., C.A.R.; Software: A.X.H.; Validation: A.X.H., E.P., B.E.N., C.A.R.; Formal analysis: A.X.H., B.E.N., C.A.R.; Investigation: A.X.H., B.E.N., C.A.R. J.A.S., M.P., S.B., M.V.K.; Resources: B.E.N., C.A.R., A.T., E.H., S.C., B.R.; Data curation: A.X.H., B.E.N., C.A.R.; Visualization: A.X.H., E.P., B.E.N., C.A.R.; Supervision: B.E.N., C.A.R.; Project administration: B.E.N., C.A.R.; Funding acquisition: B.E.N., C.A.R., A.T., E.H., S.C., B.R.; Writing – original draft: A.X.H., B.E.N., C.A.R.; Writing – review & editing: All authors.

## Declaration of interests

A.T., E.H., S.C., B.R. and B.E.N. declare that they are employed by FIND, the global alliance for diagnostics. J.A.S., M.D.P., S.B., M.V.K: The findings and conclusions in this manuscript are those of the authors and do not represent the official position of the World Health Organization.

## Notes

### Summary of Updates

Retitled, reformatted and revised for re-submission.

