## Supplementary Information for "SARS-CoV-2 diagnostic testing rates determine the sensitivity of genomic surveillance programs"

**Supplementary Notes**

Technical details of the PATAT simulation model

The computational flow of a PATAT simulation is summarized as follows: First, an age-structured population of agents is created. Close contact networks are subsequently created based on the given demographic data. The simulation is then initialized and iterates over a given period of time where each time step corresponds to a day. The sequential operations during each timestep follow the following order: (1) update the disease progression of infected individuals, (2) update the status of isolated/quarantined agents, (3) application of community testing strategies and (4) computation of transmission events within contact networks.

*Population demography*

Using input demographic data which includes information such as population age and sex distribution, household composition, employment and schooling rates, PATAT generates a population of individuals who are linked by a series of underlying contact network settings where transmission may occur. These contact network settings include households, schools, workplaces, regular mass gatherings (i.e. church) as well as random community contacts.

*Household*

PATAT randomly generates a Poisson distribution of household sizes based on the given mean household size. A reference individual (e.g. head of the household) above an assumed prime adult age (e.g. 20 years) is first randomly assigned to each household. To account for multigenerational households, the remaining household members are then randomly sampled multinomially by the input age distribution of households. Although PATAT does not explicitly model the geolocation of agents, households are ordered to implicitly approximate neighbourhood proximity.

*Schools*

PATAT distinguishes between elementary and secondary schools. For each education level, schooling children are randomly sampled from the population based on given enrolment rates and gender parity. Class sizes are then randomly drawn from a Poisson distribution based on the input mean class size while constrained by the number of schooling children attending the same grade (i.e. age; a class include only students studying the same grade). Schools are created by random allotment of classes such that (i) all schools will have equitable distributions of classes of all grades for the given education level and (ii) the total number of students approximately equals to the expected school size. Classes are then populated by schooling agents such that (i) agents of proximally ordered households will tend to attend the same school and (ii) children of the same grade (age) from identical households will not be assigned to the same class even though they may attend the same school. School teachers are then randomly drawn from the employed prime adult population based on the input teacher-to-student ratio and are assumed to have contact with each other during school days. Each class is randomly assigned to one teacher.

*Workplaces*

PATAT generates both formal and informal workplace contact networks based on separate employment rates. Youth (15-19 years) employment is also considered in the potential workforce. The distinction between formal and informal settings is made as mean employee contact rates likely differ between them. Furthermore, workplace distribution of Ag-RDTs for community testing is assumed to be feasible for formal employment entities only. Unlike schools, PATAT does not explicitly model for workplaces but sets up contact matrices between employed individuals who would be in regular contact at work. Different sizes of workplace contact networks are randomly drawn from a Poisson distribution based on the given mean employee contact size. An employed agent would only be associated with one workplace contact network.

*Mass gatherings (Churches)*

High-density mass gatherings are considered in the model in the form of contacts among church congregations given the large weekly worship attendance in Zambia (i.e. >70%)^1^ which we had modelled as our prototypical low-income country. The size of a church is assumed to follow a Normal distribution with the given mean and variance. PATAT assumes that all members of a household will visit a church together every Sunday. Other than close contacts with each other, each household member would also have a random number of close contacts from other households that attend the same church. This random contact number is drawn from a Gamma distribution with the given shape and scale parameters. Churches are also ordered such that proximally ordered households in the same neighbourhood would visit the same church.

*Random community*

PATAT assumes that every agent within a given age range would have a random number of contacts with the community daily, drawn from a Poisson distribution with a given mean.

*Disease progression*

PATAT implements a SEIRD epidemic model where the simulated population is distinguished between five compartments: susceptible, exposed (i.e. infected but is not infectious yet; latent phase), infected (which include the presymptomatic infectious period for symptomatic agents), recovered and dead. The infected compartments are further stratified by their presented symptoms, including asymptomatic, presymptomatic, symptomatic mild or severe. All symptomatic agents will also first undergo an infectious presymptomatic period after the exposed latent period. They will either develop mild symptoms who will always recover from the disease or experience severe infection which could either lead to death or recovery. PATAT uses age-structured wild-type SARS-CoV-2 disease severity and mortality probabilities as tabulated in Table S1. As a simplification, PATAT currently assumes that all agents presenting severe symptoms will be hospitalized and removed from the population.

The total duration of infection since exposure depends on the symptoms presented by the patient and is comprised of different phases (i.e. latent, asymptomatic, presymptomatic, onset-to-recovery/death) (Table S1).

*Within-host viral dynamics*

For each infected agent, PATAT explicitly simulates their viral load trajectory of cycle threshold (Ct) values over the course of their infection using a stochastic model modified from the one previously developed by ^2^. A baseline Ct value ($Ct_{baseline}$) of 40 is established upon exposure. The infected agent becomes infectious upon the end of the latent period and their Ct value is assumed to be $\leq30$. A peak Ct value is then randomly drawn from a normal distribution with the given mean and standard deviation values of the transmitted variant virus (Table S1). Peak Ct is assumed to occur upon symptom onset for symptomatic agents and one day after the latent period for asymptomatic individuals. Cessation of viral shedding (i.e. return to $Ct_{baseline}$) occurs upon recovery or death. PATAT assumes that the transition rate towards peak Ct value should not be drastically different to that when returning to baseline upon cessation (i.e. there should be no sharp increase to baseline Ct value after gradual decrease to peak Ct value or vice versa). As such, the time periods of the different phases of infection are randomly drawn from the same quintile of their respective sample distribution. The viral load trajectory is then simulated by fitting a cubic Hermite spline to the generated exposed ($t_{exposed}$, $Ct_{baseline}$), latent ($t_{latent}$, $Ct_{latent}=30$), peak ($t_{peak}$, $Ct_{peak}$) and cessation values ($t_{recovered/death}$, $Ct_{baseline}$). The slope of the fitted curve is assumed to be zero for all of them except during $t_{latent}$where its slope is assumed to be $\frac{Ct_{peak}-Ct_{baseline}}{t_{peak}-t_{exposed}}$. PATAT then uses the fitted trajectory to linearly interpolate the viral load transmissibility factor ($f_{load,i}$) of an infectious agent $i$ assuming that they are twice as transmissible at peak Ct value (i.e. $f_{load}=2$) relative to when they first become infectious (i.e. Ct value = 30; $f_{load}=1$).

*Transmissions*

When an infectious agent $i$ comes into contact with a susceptible individual $j$, the probability of transmission ($p_{transmission, (i,j)}$) is given by following equation from ^3^:

$$p_{transmission,(i,j)}=\beta\times\Phi_{i}\times f_{c}\times f_{asymp,i}\times f_{load,i}\times f_{immunity,j}\times f_{susceptiblity,j}\times\rho_{i}\times\rho_{j}$$

where $\beta$ is the base transmission probability per contact, $\Phi_{i}$ is the overdispersion factor modelling individual-level variation in secondary transmissions (i.e. superspreading events), $f_{c}$ is a relative weight adjusting $\beta$ for the network setting $c$ where the contact has occurred, $f_{asymp,i}$ is the assumed relative transmissibility factor if infector $i$ is asymptomatic, $f_{immunity,j}$ measures the immunity level of susceptible $j$ against the transmitted virus (i.e. $f_{immunity,j}=1$ if completely naïve; $f_{immunity,j}=0$ if fully protected), $f_{susceptiblity,j}$ is the age-dependent susceptibility of $j$, $\rho_{i}$ and $\rho_{j}$ are the contact rates of infector $i$ and susceptible $j$ respectively.

$\Phi_{i}$is randomly drawn from a negative binomial distribution with mean of 1.0 and shape parameter of 0.45^4^. As evidence have been mixed as to whether asymptomatic agents are less transmissible, we conservatively assume there is no difference relative to symptomatic patients (i.e. $f_{asymp,i}=1$). The age-structured relative susceptibility values $f_{susceptiblity,j}$ are derived from odds ratios reported by ^5^ (Table S1).

$\beta$ is determined by running initial test simulations with a range of values on a naïve population with no interventions that would satisfy the target basic reproduction number $R_{0}$ as computed from the resulting exponential growth rate and distribution of generation intervals ^6^. $f_{c}$ is similarly calibrated during these test runs such that the transmission probabilities in households, workplaces, schools, and all other community contacts are constrained by a relative weighting of 10:2:2:1 ^3^.

*Testing by Ag-RDT*

Unlike PCR which is highly sensitive due to prior amplification of viral genetic materials, the sensitivity of Ag-RDT depends on the viral load of the tested patient. While the specificity of Ag-RDT is assumed to be 98.9%, its sensitivity depends on the Ct values of the tested infected agent: Ct $>35$ (0%); 35 – 30 (20.9%); 29 – 25 (50.7%); Ct $\leq24$ (95.8%) ^7^.

Testing by Ag-RDT may either occur via symptomatic testing at healthcare facilities. First, a symptomatic agent may opt to go into self-isolation upon symptom onset prior to being tested, as decided by a Bernoulli trial with probability $p_{self-isolation}$. Regardless if they were self-isolated, after $\tau_{delay,symp-test}$ days from symptom onset, the symptomatic agent may then decide to get tested with a Bernoulli probability of $p_{symp-test}$ that inversely correlates with the distance between the agent’s household and the nearest healthcare facility (Table S1). PATAT assumes that agents who have decided against symptomatic testing (i.e. failed Bernoulli trial) or received negative test results will not seek symptomatic testing again.

*Isolation and quarantine*

We assumed that agents would change their behaviour when (i) they start to present symptoms and go into self-isolation (10% compliance assumed, 71% endpoint adherence ^8^); (ii) they test positive and are isolated for 10 days (50% compliance assumed, 86% endpoint adherence ^8^); or (iii) they are household members (without symptoms) of positively-tested agents and are required to be in quarantine for 14 days (50% compliance assumed, 28% endpoint adherence ^8^). Once an agent goes into isolation/quarantine, we linearly interpolate their probability of adherence to stay in isolation/quarantine over the respective period. Given the lack of infrastructure and resources to set up dedicated isolation/quarantine facilities in many low-middle income countries, we assumed that all isolated and quarantined individuals would do so at home. Although they have no contact with agents outside of their home, we assumed that they would maintain 90% contact rate with household members.

Background on current guidance

Here, we provide relevant details on the mathematical frameworks underlying three current guidance on minimum sequencing samples required for variant detection mentioned in the main text. Specifically, we highlighted the critical assumptions made and the lack of consideration of spatiotemporal bias resulting from low testing volumes and sampling coverage in each approach.

The World Health Organization (WHO) and European Centre for Disease Prevention and Control (ECDC) computes sequencing sample size using the binomial method.^9,10^ **Binomial sampling assumes that specimens to be sampled collected for sequencing are randomly representative of the circulating virus diversity**. As acknowledged by the WHO and ECDC, this is difficult to achieve with low testing rates and spatial non-uniformity in sampling coverage which can introduce spatiotemporal biases in sequencing samples. However, there was no advice in the guidance on how to correct for these biases.

Brito et al. made recommendations on sequencing sample size by computing the probability of detecting at least one variant genome under different sequencing proportions of detected cases based on random subsampling of genomic surveillance data collected in Denmark in 2020-2021^11^. Data from Denmark was used as it was one of the most comprehensive genomic surveillance programs in the world – they were sequencing at >10% of detected cases in most weeks. Brito et al. estimated that sequencing 0.5% of all detected cases would result in sequencing at least one variant genome before the number of variant infections reach 100 cases if turnaround time is kept at 21 days. However, there would only be a 20% probability that this would occur based on their subsampling analyses. More importantly, Denmark was testing at one of the highest rates in the world during this period, performing >2,000 tests/100,000 people/day on average (<https://www.finddx.org/covid-19/test-tracker/>). **Brito et al., however, did not extend their subsampling analyses on the virus diversity among the detected cases under lower testing rates.** This would have otherwise provided corrections on the suggested sampling proportions under lower testing volumes.

Wohl et al. provided the following derivation to compute the sequencing sample size per unit time ($n$) for *ongoing* surveillance:^12^

$$n\approx-\frac{ln\left[ 1-\Pr\left( d\leq t \right) \right]}{G(t)-G(0)}$$

where $\Pr\left( d\leq t \right)$ is the probability of detection on or before time $t$ and $G(t)$ is the cumulative density function that model the growth in *circulating* variant proportion over time. Wohl et al. applied the logistic growth curve function to *circulating* variant proportion growth. As such:

$$G'(t)=\frac{1}{r}ln\left| a+e^{rt} \right|+C$$

where $r$ is the assumed per unit-time growth rate of the variant and $a=\frac{1}{p_{0}}-1$ where $p_{0}$ is the initial variant virus proportion.

**In this form, Wohl et al. assumes that the *observed* variant proportion in the positive specimens collected perfectly matches the *circulating* variant proportion among the infected population. This is only possible if testing volumes are sufficiently large enough.** At the proposed target of 1% *circulating* variant proportion, at least 38,031 tests must be performed in total each day to ensure that the *observed* variant proportion is also at 1% with a margin error of 0.1% at 95% confidence. For ~18 million people in Zambia, this means that the average testing rate be maintained at 212 tests/100,000 people/day. This is ~8 times more than the average LMIC testing rate of 27 tests/100,000 people/day.

To account for likely enriched *observed* variant proportion in the sample pool sent for sequencing, Wohl et al incorporated a correction factor $\frac{C_{Ext}}{C_{Var}}$ to the *circulating* variant proportion such that:

$$G'(t)=\frac{1}{r}ln\left| \left( \frac{C_{Ext}}{C_{Var}} \right)a+e^{rt} \right|+C$$

$\frac{C_{Ext}}{C_{Var}}$ is the ratio of the coefficient of detection for the extant ($C_{Ext}$) over that for the variant virus ($C_{Var}$). $C_{Ext}$ and $C_{Var}$ are essentially the joint conditional probabilities of obtaining a variant and extant virus sequence respectively. For the virus $v$, the coefficient of detection ($C_{v}$) is defined as:

$$C_{v}=\phi_{v}\gamma_{v}\left[ \alpha_{v}\beta_{a}+\left( 1-\alpha_{v} \right)\beta_{s} \right]$$

where $\phi_{v}$ is the sensitivity of the diagnostic test to virus $v$, $\gamma_{v}$ is the probability that the detected infection caused by virus $v$ meets the quality threshold for sequencing (i.e. below the stipulated PCR cycle threshold value), $\alpha_{v}$ is the probability a person infected with virus $v$ is asymptomatic, $\beta_{a}$ and $\beta_{s}$ are the probabilities an asymptomatic and symptomatic person infected with SARS-CoV-2 were tested (regardless of which variant they were infected by).

Although $\beta_{a}$ and $\beta_{s}$ are incorporated in $\frac{C_{Ext}}{C_{Var}}$, the correction factor computes the relative likelihood of detection between the extant and variant virus. In other words, $\frac{\boldsymbol{C}_{\boldsymbol{Ext}}}{\boldsymbol{C}_{\boldsymbol{Var}}}$ **only corrects for biases in the *observed* variant proportion due to relative differences in diagnostic sensitivities, sample qualities and conditional asymptomatic and symptomatic testing probabilities between the two viral variants.** **It does not factor in distortions in the *observed* variant proportion from other sources of bias, including: (1) stochastic effects arising from low testing volumes and unevenness of daily test stock availability, (2) unconditional probability of asymptomatic testing, and (3) spatial biases when only a subset of samples from sentinel sites are sent for sequencing.**

Emergence of SARS-CoV-2 variants of concern

Here, we briefly recount the emergence of other SARS-CoV-2 variants-of-concern (VOC; i.e. the Alpha, Beta, Gamma and Delta) besides Omicron (i.e. timepoint in collection of the first variant sequence) given the prevailing circumstance, then-level of testing and sequencing performed in the respective countries where they likely first emerged from.

*Alpha variant*

The Alpha variant was first reported in the UK in early December 2020 after public health agencies investigated the rapid increase in COVID-19 cases in Kent, South East England despite prevailing high levels of non-pharmaceutical interventions ^13^. Retrospective analyses found that the first Alpha virus that was later sequenced was collected on 20 September 2020 and phylogenetic analyses estimated the time to most recent common ancestor (TMRCA) of the Alpha lineage to be around the same time^14^. The UK was testing at a mean rate of >300 tests per 100,000 people per day (tests/100k/day) in September 2020 ^15^ and randomly sampling an average of 7.9% cases every week in Kent for sequencing ^14^. From on our genomic surveillance simulation results, albeit derived for a Zambian population, we would expect the first Alpha virus variant to be collected for sequencing within one week of its emergence under a random sampling approach at these testing and sequencing rates.

*Beta variant*

The Beta variant was first reported in South Africa in December 2020 as the country experienced its second wave of SARS-CoV-2 infections. The earliest Beta virus selected for sequencing was collected on 8 October 2020 and the TMRCA was estimated to between July and August 2020 ^16^. South Africa was still in the midst of the peak of the first wave of infection during the estimated TMRCA period and testing at mean rates of ~60 and ~40 tests/100k/day in July and August 2020 respectively ^15^. Only ~0.3% of cases identified in South Africa in July-August 2020 were sequenced and deposited in the GISAID EpiCoV database ^17,18^. The estimated ~2-3-month delay in sampling the first Beta variant sequence is therefore likely based on our simulations due to a combination of relatively low levels of testing and sequencing that was further exacerbated by the variant virus emerging during the peak circulation of the extant SARS-CoV-2 wild-type virus.

*Gamma variant*

The Gamma variant was first reported in January 2021 in Brazil as a result of investigating the rapid rise in hospitalizations in Manaus in December 2020 ^19^ as well as in Japan from infected travelers who recently returned from the Amazonas ^20^. The first Gamma variant virus selected for sequencing was collected on 6 December 2020 and phylogenetic analyses estimated that the VOC lineage likely emerged in Manaus between October and November 2020 ^19^. During this period, Brazil was testing at 10-30 tests/100k/day on average ^15^ and sequenced ~0.1% of all confirmed cases ^17,18^. By early January 2021 when the sequencing results were obtained and shared, the circulating proportion of the Gamma variant in Manaus was estimated to be ~75% ^19^. The 1-2 month gap between emergence and sampling of a VOC sequence is likely due to low testing and sequencing rates, consistent with the results of our simulations. Moreover, the turnaround time between sample collection and sequencing data acquisition added an additional month in delay before the Gamma variant was first reported in Brazil.

*Delta variant*

The earliest Delta (i.e. PANGO lineage B.1.617.2) sequence (Accession: EPI_ISL_9232357) collected in India that is deposited in the GISAID EpiCoV database was collected on 3 September 2020. This sample however was only sequenced retrospectively as (i) it was submitted to the database on 28 January 2022 and (ii) published works by the Indian SARS-CoV-2 Genomics Consortium (INSACOG), the national sentinel sequencing network, referred to earliest identification of Delta in state of Maharashtra in December 2020 ^21,22^. The identification of Delta in Maharashtra was only done so retrospectively to investigate the surge in cases in the state in January 2021. Using Delta sequences collected globally, the likely TMRCA period was estimated to be around September 2020 as well ^23^. During this time, India was still experiencing the peak of its first wave of SARS-CoV-2 infections ^18^. The second wave of SARS-CoV-2 infections across the country caused by the Delta variant only took off in March 2021, six months after the estimated TMRCA ^24^. Between December 2020 when the first wave of infections subsided and the beginning of the second wave in March 2021, there were several competing lineages circulating in India, including the Alpha VOC as well as the Kappa variant of interest (i.e. B.1.617.1), a sub-lineage descending from the same parental lineage of Delta (i.e. B.1.617) ^24^. There were no coordinated efforts to perform active genomic surveillance across India in 2020; Sequencing analyses then were largely performed retrospectively in response to surge in cases ^22,25^. The INSACOG was only established by the government on 30 December 2020 in response to monitor genetic variations in light of the introduction of the Alpha variant into the country ^26,27^. Owing to complexities attributed to multiple co-circulating and competing variant lineages, nonuniformity in sampling, in part due to a lack of ongoing active coordinated nation-wide genomic surveillance efforts then, and that the “earliest” Indian Delta sequences were all identified from retrospective analyses, there are uncertainties around both the emergence and early spread of the Delta variant within India.

**Extended Data Figures**

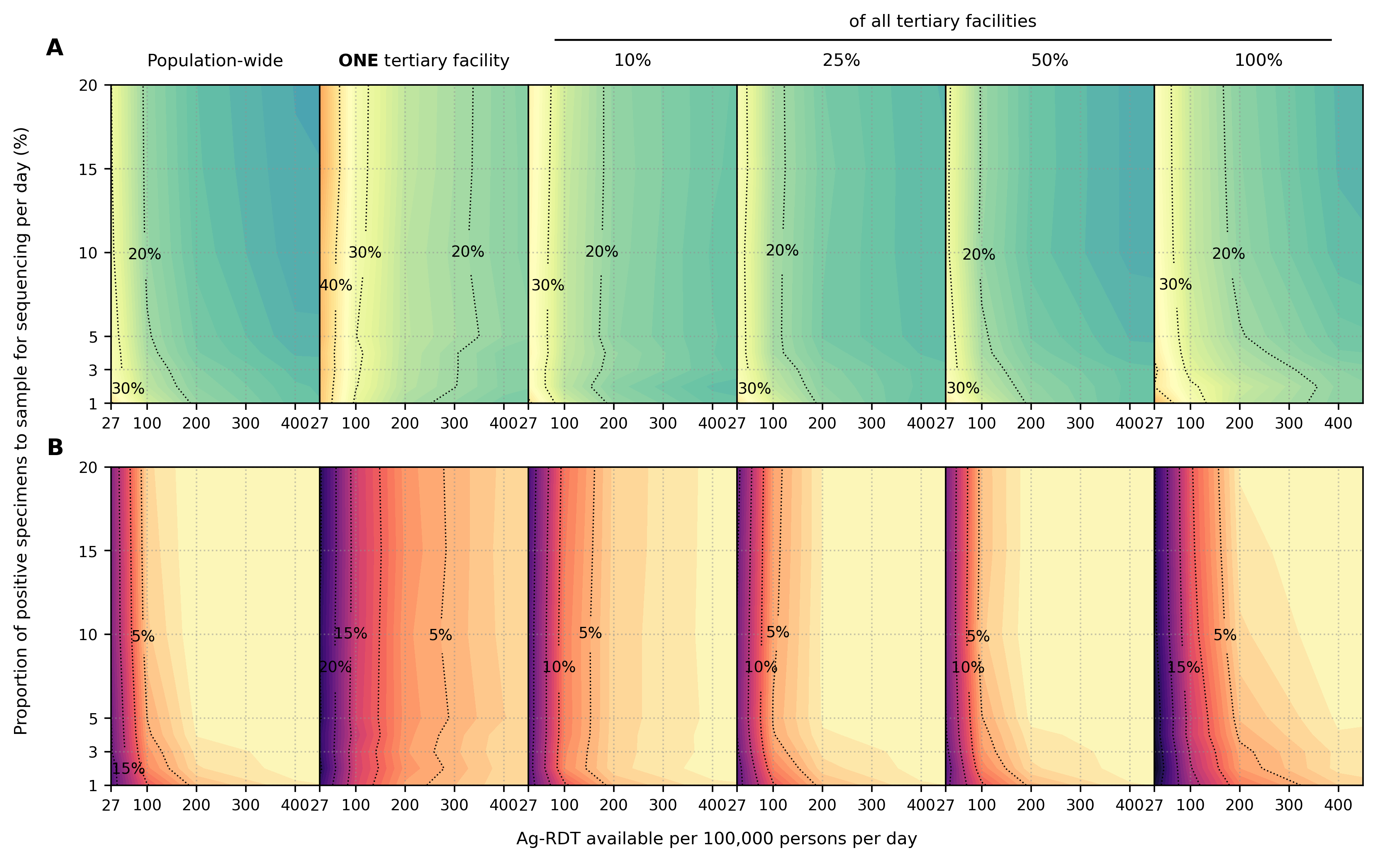

**Extended Data Fig. S1. Impact of SARS-CoV-2 Ag-RDT testing rates and daily proportion of positive specimens to sample for sequencing on observed Omicron variant proportions**. Different genomic surveillance strategies (i.e. all specimens collected from all healthcare facilities sent to one facility to be sampled for sequencing (*population-wide* strategy); only *one*, 10%, 25%, 50% or 100% of all tertiary facilities acting as sentinel sites that would sample the specimens they collected for sequencing) were simulated. (**A**) Maximum absolute difference between observed and circulating variant proportions. (**B**) Proportion of timepoints when sequencing was performed that the absolute difference between observed and circulating variant proportions is greater than 20%. All results were computed from 1,000 random independent simulations for each surveillance strategy.

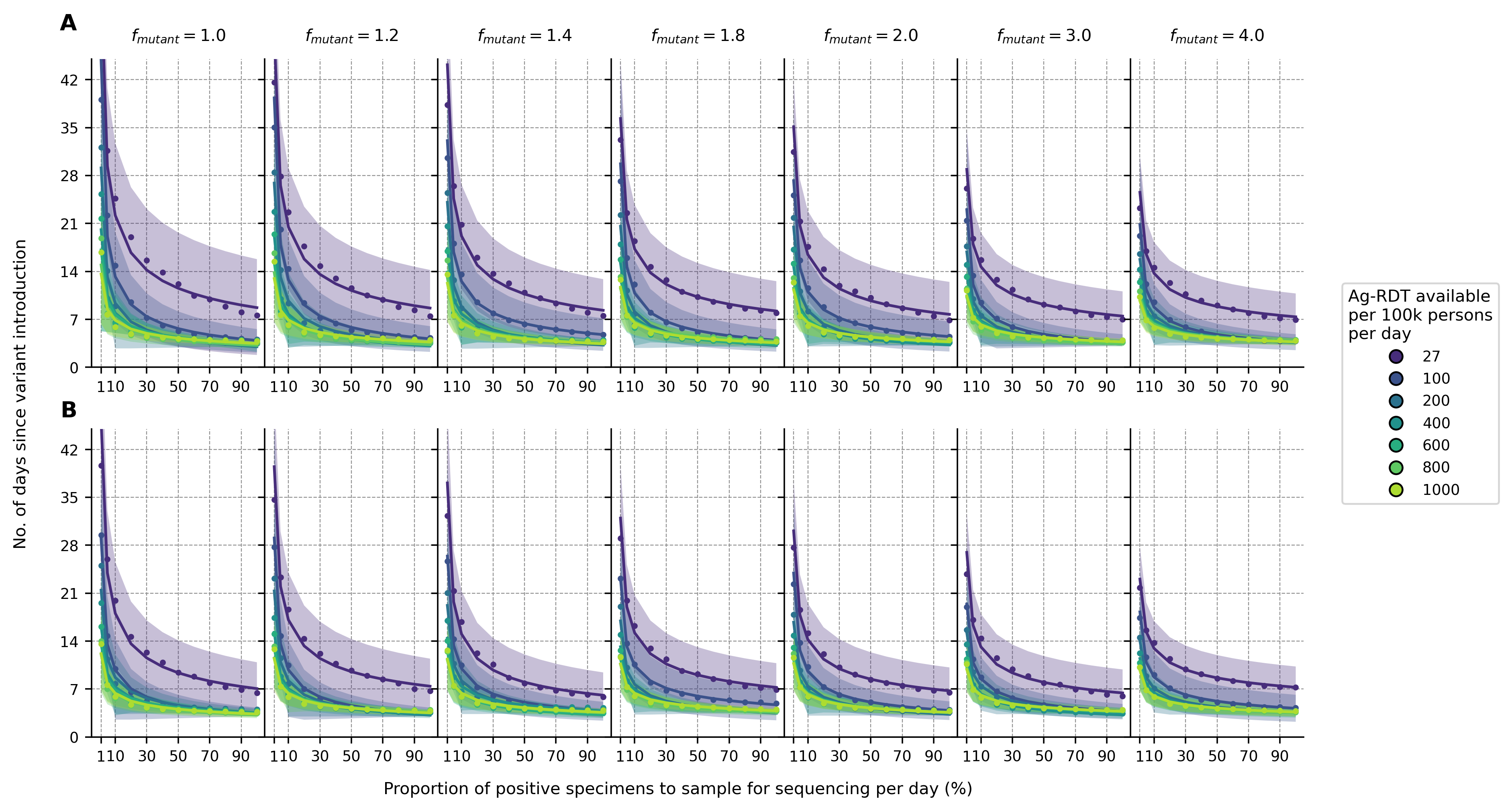

**Extended Data Fig. S2. Sensitivity analyses on variant detection operating curve for different relative transmissibility factor**. For each Ag-RDT availability, the expected day when the first Omicron variant specimen (in the background of extant Delta variant) is sampled for sequencing since its introduction is plotted against the proportion of positive specimens to be sampled for sequencing daily. All specimens collected from the population from all healthcare facilities were sent to one facility to be sampled for sequencing (population-wide genomic surveillance strategy). Different transmissibility factor of Omicron relative to Delta ($f_{mutant}$) were assumed. (**A**) 10% and (**B**) 40% of the population had immunity against Omicron initially. All results were computed from 1,000 random independent simulations for each surveillance strategy. The shaded region depicts the standard deviation across simulations.

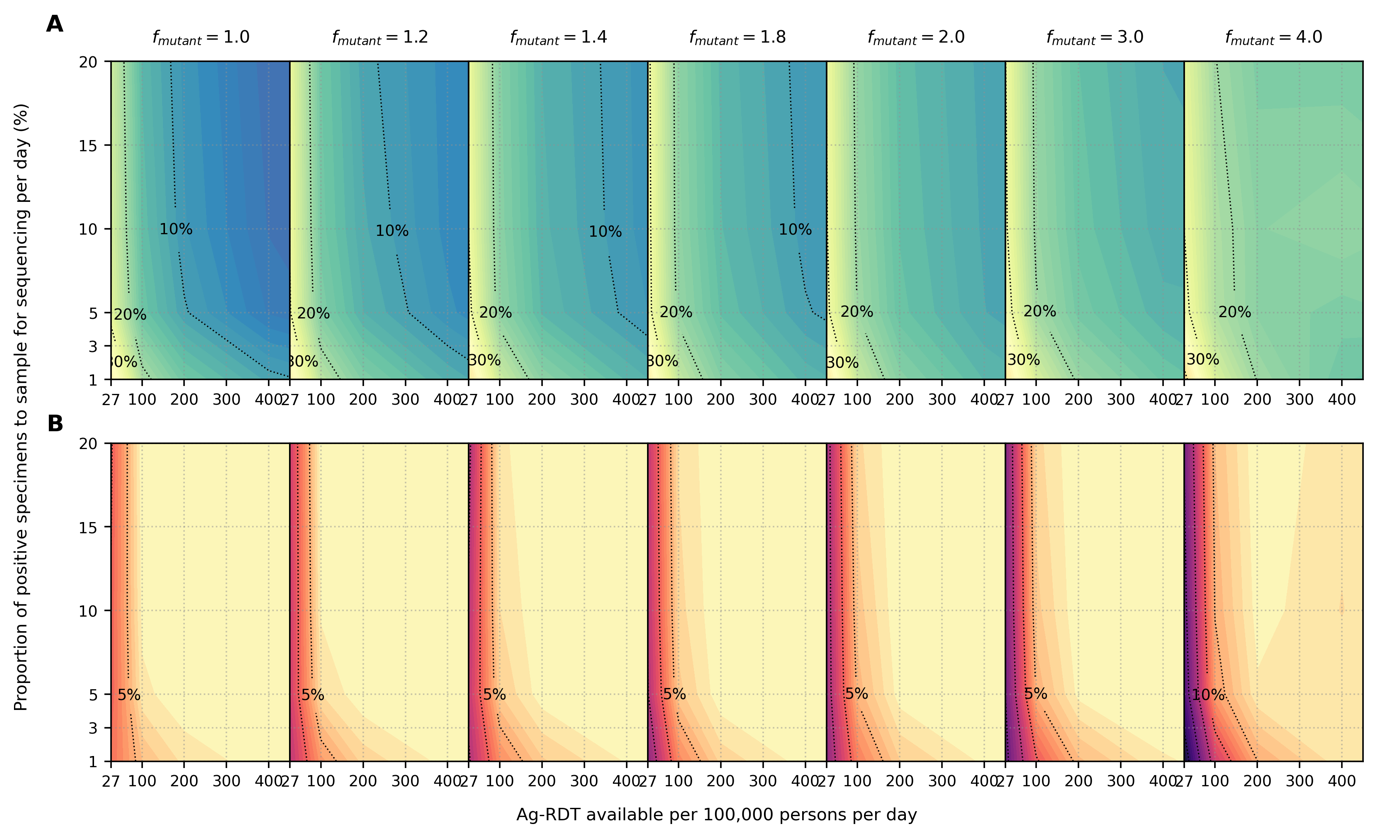

**Extended Data Fig. S3. Sensitivity analyses on accuracy of observed variant proportions for different relative transmissibility factor**. Omicron-like virus properties assumed for variant and initial proportion of population with some degree of protection against the variant virus assumed at 10%. All specimens collected from the population from all healthcare facilities were sent to one facility to be sampled for sequencing (population-wide genomic surveillance strategy). Different transmissibility factor of Omicron relative to Delta ($f_{mutant}$) were assumed. (**A**) Maximum absolute difference between observed and circulating variant proportions. (**B**) Proportion of timepoints when sequencing was performed that the absolute difference between observed and circulating variant proportions is greater than 20%. All results were computed from 1,000 random independent simulations for each surveillance strategy.

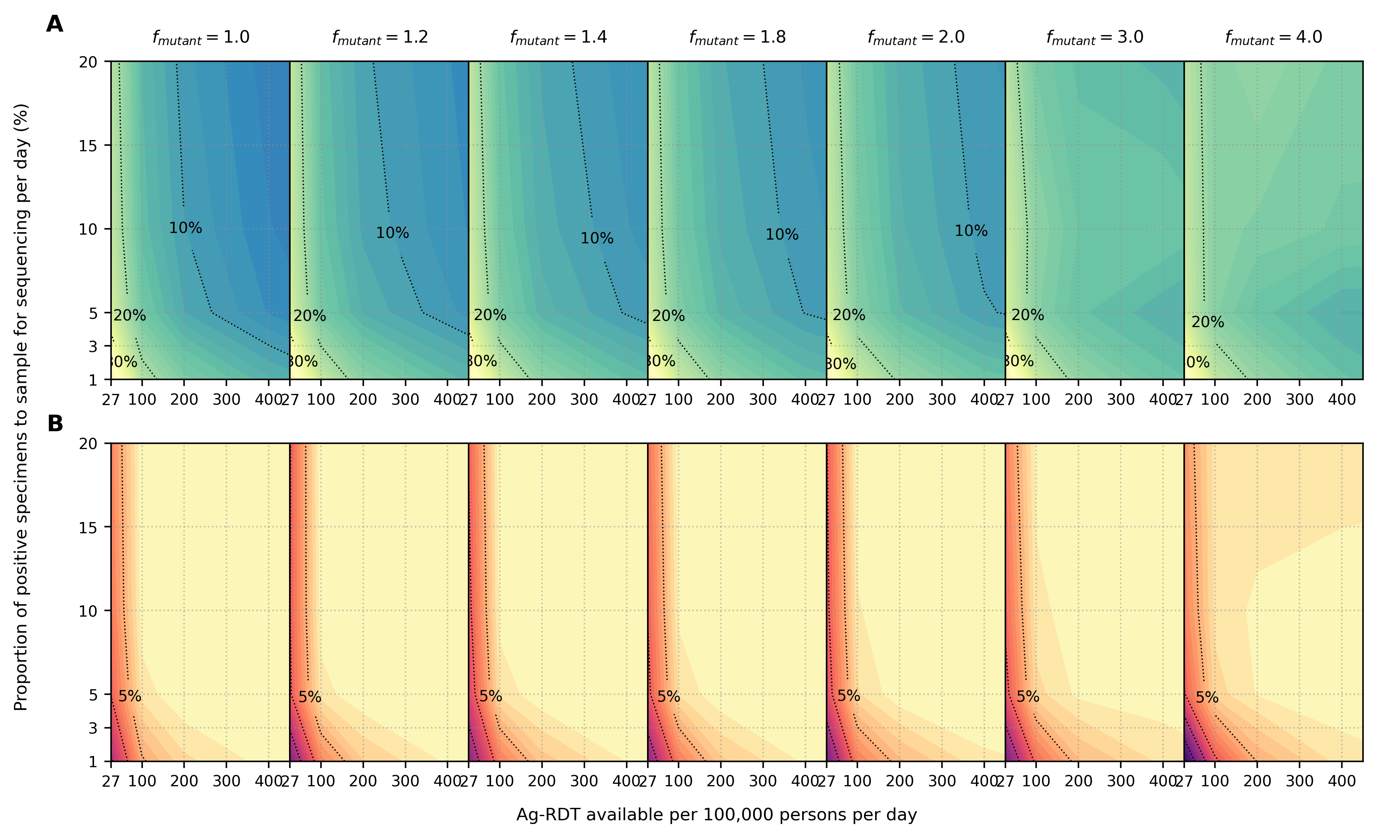

**Extended Data Fig. S4. Sensitivity analyses on accuracy of observed variant proportions for different relative transmissibility factor**. Omicron-like virus properties assumed for variant and initial proportion of population with some degree of protection against the variant virus assumed at 40%. All specimens collected from the population from all healthcare facilities were sent to one facility to be sampled for sequencing (population-wide genomic surveillance strategy). Different transmissibility factor of Omicron relative to Delta ($f_{mutant}$) were assumed. (**A**) Maximum absolute difference between observed and circulating variant proportions. (**B**) Proportion of timepoints when sequencing was performed that the absolute difference between observed and circulating variant proportions is greater than 20%. All results were computed from 1,000 random independent simulations for each surveillance strategy.

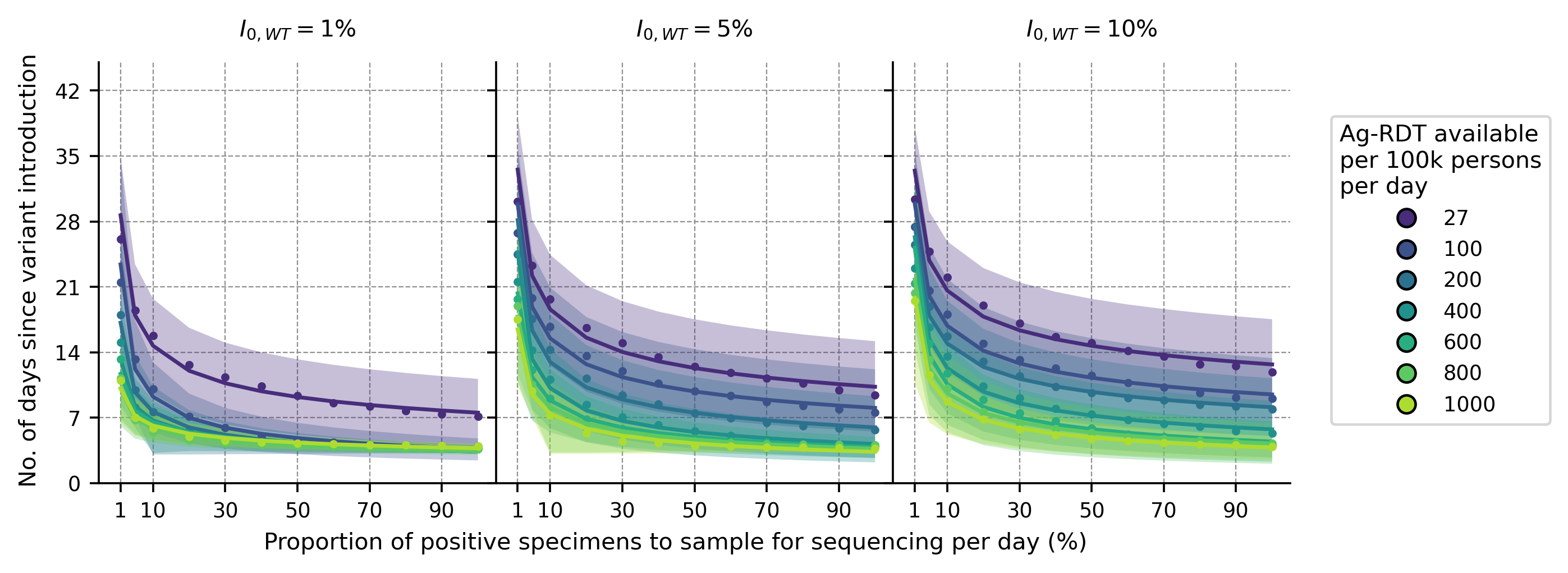

**Extended Data Fig. S5. Impact of prevalence of extant variant of concern (**$\boldsymbol{I}_{\mathbf{0,}\boldsymbol{WT}}$**) at the time of new variant introduction**. For each Ag-RDT availability, the expected day when the first Omicron variant specimen (in the background of Delta) is sampled for sequencing since its introduction is plotted against the proportion of positive specimens to be sampled for sequencing daily. Each panel shows a different prevalence of the Delta variant ($I_{0,WT}$) at the point of Omicron introduction. Sampling for sequencing was drawn from the population-wide scenario. All results were computed from 1,000 random independent simulations for each surveillance strategy. The shaded region depicts the standard deviation across simulations.

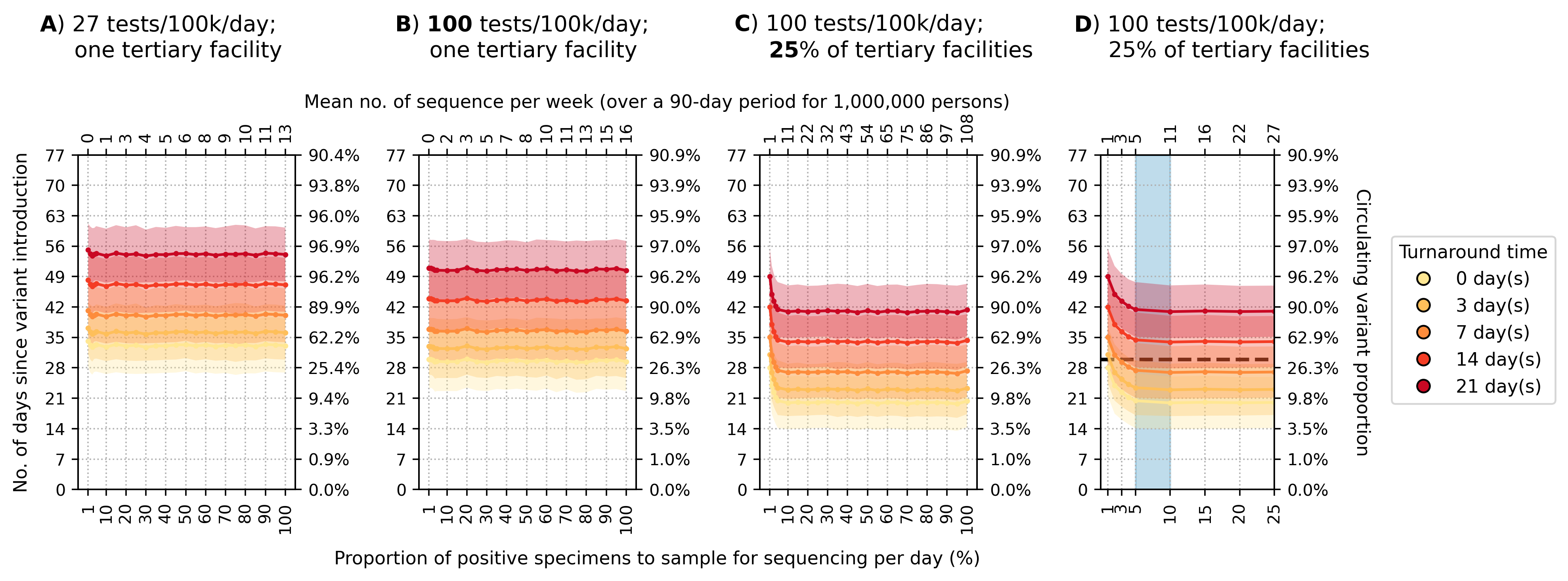

**Extended Data Fig. S6. Recommended approach to enhance genomic surveillance robustness**. In each plot, the operating curves of the expected day when the first Omicron BA.1 variant sequence is generated are plotted for different proportion of specimens to sample for sequencing per day and turnaround times. We assumed that the Omicron BA.1 variant was circulating at 1% initially with Delta variant in the background. We also assumed that positive specimens sampled within each week for sequencing are consolidated into a batch before they are referred for sequencing. Turnaround time refers to the time between collection of each weekly consolidated batch of positive specimens to the acquisition of its corresponding sequencing data. The vertical axes denote the number of days passed since the introduction of the Omicron variant (left) and its corresponding circulating proportion (right). The horizontal axes denote the proportion of positive specimens to sample for sequencing per day (bottom) and the corresponding mean number of sequences to be generated per week per 1,000,000 people over a 90-day epidemic period. (**A**) Specimen pools for sequencing from *one* tertiary facility with testing rate at 27 tests per 100,000 persons per day (tests/100k/day). (**B**) Specimen pools for sequencing from *one* tertiary sentinel facility with testing rate at 100 tests/100k/day. (**C**) Specimen pools for sequencing from 25% of all tertiary facilities acting as sentinel sites with testing rate at 100 tests/100k/day. (**D**) Zoomed-in plot of (C) for sequencing proportions varying between 1-25%. Sequencing 5-10% of positive specimens (blue shaded region) would ensure that we would expectedly detect Omicron within 30 days if turnaround time is kept within one week. All results were computed from 1,000 random independent simulations for each surveillance strategy. The shaded region depicts the standard deviation across simulations.

**Extended Data Tables**

**Extended Data Table S1. PATAT simulation parameters**.

| Parameter | Values/Distribution | Source |
| --- | --- | --- |
| *Population demography* | | |
| Total population size | 1,000,000 |  |
| Mean household size | 5.0 | ^28^ |
| Age structure (in bins of 5 years) | [0.161, 0.165, 0.157, 0.101, 0.083, 0.068, 0.057, 0.051, 0.042, 0.030, 0.024, 0.015, 0.016, 0.009, 0.008, 0.005, 0.006, 0.002, 0.000, 0.000] | ^28^ |
| Minimum prime adult age | 20 years | Assumed |
| Proportion of women | 51% | ^29^ |
| Minimum working age | 15 years | ^29^ |
| Employment rate | 39% (male), 23% (female) | ^29^ |
| Formal employment rate | 36% (employed male), 24% (employed female) | ^29^ |
| Schooling rate | 79% (male), 40% (female) | ^28^ |
| School gender parity | 1.0 (Primary), 0.9 (Secondary) | ^28^ |
| Church participation rate | 70% of all households | Assumed |
| Mean employment contacts (formal) | 20 | Assumed |
| Mean employment contacts (informal) | 5 | Assumed |
| Mean class size | 37 (Primary and secondary) | ^28^ |
| Mean school size | 700 (Primary and secondary) | Assumed |
| Student-teacher ratio | 42 (Primary and secondary) | ^28^ |
| Mean church size (s.d.) | 500 (100) | Assumed |
| Mean random contacts in church per person | 10 | Assumed |
| Mean random community contacts per day | 10 | Assumed |
| *SARS-CoV-2 transmissions related parameters* | | |
| Age-structured relative susceptibility (in bins of 5 years) | [0.34, 0.34, 0.67, 0.67, 1.00, 1.00, 1.00, 1.00, 1.00, 1.00, 1.00, 1.00, 1.00, 1.00, 1.24, 1.24, 1.47, 1.47, 1.47, 1.47] | ^3,5^ |
| Age-structured probability of becoming symptomatic (in bins of 5 years) | [0.50, 0.50, 0.55, 0.55, 0.60, 0.60, 0.65, 0.65, 0.70, 0.70, 0.75, 0.75, 0.80, 0.80, 0.85, 0.85, 0.90, 0.90, 0.90, 0.90] | ^30,31^ |
| Age-structured probability of developing severe disease (in bins of 5 years) | [0.00050, 0.00050, 0.00165, 0.00165, 0.00720, 0.00720, 0.02080, 0.02080, 0.03430, 0.03430, 0.07650, 0.07650, 0.13280, 0.13280, 0.20655, 0.20655, 0.24570, 0.24570, 0.24570, 0.24570] | ^30,31^ |
| Age-structured probability of death (in bins of 5 years) | [0.00002, 0.00002, 0.00002, 0.00002, 0.00010, 0.00010, 0.00032, 0.00032, 0.00098, 0.00098, 0.00265, 0.00265, 0.00766, 0.00766, 0.02439, 0.02439, 0.08292, 0.08292, 0.16190, 0.16190] | ^8,32^ |
| Latent period (days) | Wild-type SARS-CoV-2/Alpha: Lognormal (4.5, 1.5)  Delta/Omicron: Lognormal (4.0, 1.3) | ^3,33–35^ |
| Pre-symptomatic period (days) | Wild-type SARS-CoV-2/Alpha: Lognormal (1.1, 0.9)  Delta/Omicron: Lognormal (1.8, 1.7) | ^3,33,35^ |
| Period between symptom onset and severe disease (days) | Lognormal (6.6, 4.9) | ^33^ |
| Period between severe disease and death (days) | Lognormal (8.6, 6.7) | ^33^ |
| Recovery period for symptomatic agents with mild disease (days) | Wild-type SARS-CoV-2/Alpha: Lognormal (8.0, 2.0)  Delta: Lognormal (6.23, 0.53^*^)  Omicron: Lognormal (5.35, 0.37^*^) | ^35,36^ |
| Recovery period for asymptomatic agent (days) | Wild-type SARS-CoV-2/Alpha: Lognormal (8.0, 2.0)  Delta: Lognormal (6.23, 0.53^*^)  Omicron: Lognormal (5.35, 0.37^*^) | ^35,36^ |
| Recovery period of agents with severe disease (days) | Lognormal (18.1, 6.3) | ^30^ |
| Peak Ct values | Wild-type SARS-CoV-2/Alpha/Delta: Normal (20.5, 0.79^*^)  Omicron: Normal (23.3, 0.58^*^) | ^35^ |
| Cross-immunity to variant virus after infection by extant virus | Wild-type SARS-CoV-2/Alpha: 87%  Delta/Omicron: 20% | ^37,38^ |
| Severity (chance of hospitalization) of variant relative to extant virus | Wild-type SARS-CoV-2/Alpha: 100%  Delta/Omicron: 40% | ^39^ |
| *Testing parameters* | | |
| Delay in visiting healthcare facility for symptomatic testing (days) | Lognormal (1.0, 0.5) | Assumed |
| Ag-RDT specificity | 0.989 | ^7^ |
| Agents to healthcare facilities ratio | 7,000:1 | ^40,41^ |
| Distance-structured distribution of households to nearest healthcare facility (in bins of 1km) | [0.048, 0.193, 0.119, 0.08, 0.074, 0.098, 0.068, 0.072, 0.056, 0.191] | ^42^ |
| Distance-structured probabilities of agent visiting nearest healthcare facility for testing services (in bins of 1km) | [0.853, 0.808, 0.762, 0.717, 0.672, 0.626, 0.581, 0.536, 0.49, 0.445] | ^42^ |
| *Isolation/quarantine parameters* | | |
| Isolation period | 10 days |  |
| Quarantine period | 14 days |  |
| Self-isolation period | 10 days |  |
| Reduction in contact rates under isolation/quarantine (in order of households, schools, workplaces, church and random community) | [10%, 100%, 100%, 100%, 100%] |  |

^*^Standard deviation values inferred from 95% confidence interval computed in reference.

17.	GISAID. GISAID - Initiative. https://www.gisaid.org/.

18.	Johns Hopkins University Center for Systems Science and Engineering. CSSEGISandData/COVID-19: Novel Coronavirus (COVID-19) Cases, provided by JHU CSSE. https://github.com/CSSEGISandData/COVID-19.

27.	Ministry of Health and Family Welfare. Q&A on Indian SARS-CoV-2 Genomics Consortium (INSACOG) | Department of Biotechnology. https://dbtindia.gov.in/pressrelease/qa-indian-sars-cov-2-genomics-consortium-insacog.
